# The hospital “Sink-ome”: Pathogen and antimicrobial resistance gene burden in sink-traps across 29 UK hospitals and associations with sink characteristics

**DOI:** 10.1101/2025.07.25.25332191

**Authors:** KK Chau, G Rodger, TP Quan, E Dietz, E Pritchard, W Matlock, P Aranega Bou, G Moore, the SinkBug Consortium, A Roohi, R Hope, S Hopkins, KL Hopkins, AS Walker, N Stoesser

## Abstract

Hospital sinks are reservoirs for common healthcare-associated pathogens and antimicrobial resistance (AMR) genes, but there are limited large-scale data on the distribution of this burden across hospitals and associations with sink infrastructure-related features such as design, water chemistry, and usage. Using metagenomics to characterise the microbiome of 287 sink-traps across 29 UK hospitals, we evaluate associations between 49 sink-related features and major clinical pathogen burden, total AMR gene burden, and burden of major extended-spectrum beta-lactamase and carbapenemase genes. We show that higher clinical pathogen burden is associated with higher AMR gene burden and lower sink-trap species diversity and richness. Overgrowth of drug-resistant pathogens in these reservoirs is associated with sink-trap chemistry (including antibiotic residues, iron and alkalinity), moisture, sink location and cleaning frequency. Our study provides novel insights into the ecology of these reservoirs and identifies factors potentially amenable to intervention to reduce the risk hospital sinks pose to patients.

## Main

Healthcare-associated infections, especially those caused by Gram-negative bacilli (e.g. Enterobacterales, Pseudomonas aeruginosa) are a major cause of morbidity and mortality worldwide^1,2^. Antimicrobial resistance (AMR) is a significant global concern, with the emergence of broad-spectrum AMR mechanisms (e.g. extended-spectrum beta-lactamases, carbapenemases) rendering these infections increasingly difficult to treat^3,4^. In the UK there are marked regional differences in Gram-negative bloodstream infection rates and the prevalence of major AMR mechanisms, as highlighted in the English Surveillance Programme for Antimicrobial Utilisation and Resistance (ESPAUR) reports^2^.

Sink-traps are major nosocomial reservoirs of AMR genes and Gram-negative pathogens^5^, and dissemination of drug-resistant bacteria from sink-traps to patients may occur undetected^6^. Sink-traps also represent important niches for AMR gene exchange amongst strains and species^7^. They may therefore contribute substantially to healthcare-associated infections and AMR emergence. Several observational and interventional studies support this, with sink/drain-decontamination or removal reducing the incidence of Gram-negative infections and/or terminating AMR-associated outbreaks^8^. Nevertheless, complete removal of hospital sink infrastructure may be challenging given the importance attached to sanitary facilities in patient care and in mitigating infections such as Clostridioides difficile and norovirus.

Experimental data have shown that sink design and usage can influence bacterial transmission to patients^9^. Additionally, the presence of antibiotics and chemical composition of wastewater may affect microbiome composition and dynamics^10,11^.

However, limited real-world information on sink-trap microbiomes and the impact of sink characteristics on sink-trap pathogen and AMR burden is available across hospital settings, and this could facilitate interventions to mitigate the risk from these reservoirs (e.g. by optimising sink design/cleaning protocols), potentially without needing to remove sink infrastructure altogether.

We therefore undertook a multicentre survey of sinks and sink-traps across 29 UK hospitals (the SinkBug study) to characterise bacterial species and AMR gene diversity in sink-traps and define sink characteristics that might impact the sink-trap microbiome (“sink-ome”). We used multivariable models to investigate associations between sink and sink-trap characteristics and the sink-ome, including the abundance of major clinical pathogens and important AMR gene families, aiming to identify modifiable characteristics that could inform interventions to improve “water-safe” care.

## Results

Partnering with the National Infection Teams Collaborative for Audit and Research, we surveyed/sampled 287 sinks/sink-traps across 29 UK hospitals (18 England, 1 Northern Ireland, 6 Scotland, 4 Wales [Fig.1), January-March 2023. Sink-trap samples underwent antibiotic/chemical dipstick testing and metagenomic sequencing (see Methods). 27 sites submitted data/samples for 10 sinks/sink-traps each, one site for 9 sinks/sink-traps (site 29) and one site for 8 sinks/sink-traps (site 3). 111/287 (39%) sampled sink-traps were in intensive/high-dependency care wards (ICU), 92/287 (32%) on medical wards, and 84/287 (29%) on surgical wards. Within these wards, 97/287 (32%) sinks were in patient bays, 88/287 (31%) in sluice rooms (where patient/ward waste are discarded), 77/287 (27%) in medicines/drug preparation rooms, and 25/287 (9%) in patient side-rooms. The median sink-trap metagenome sequencing depth was 15,860,867 paired-end reads (IQR:12,670,701-18,754,114; median 4.76 Gb [IQR:3.80-5.63]). There was no evidence of association between metagenomic richness and sequencing depth (Extended Data Fig.1), suggesting that we captured the taxonomic/antimicrobial resistance gene (ARG) diversity present.

**Figure 1:**
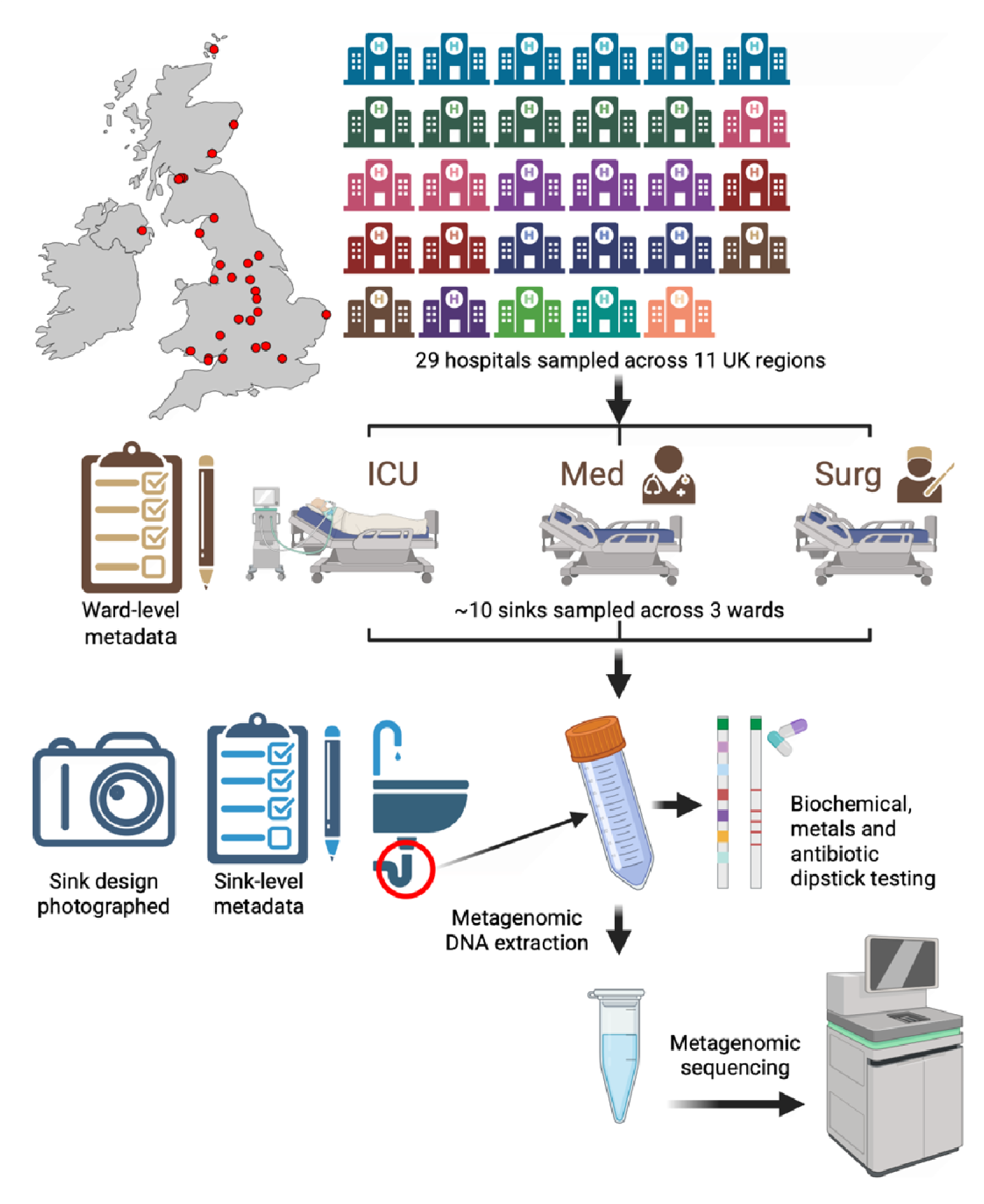
Overview of SinkBug sampling and sample processing strategy. Hospital colour reflects the region sampled. “ICU” – intensive care unit, “Med” = medical ward, “Surg” = surgical ward. Metagenomic sequencing was performed on the Illumina NovaSeq.

### Bacterial species diversity in sink-traps

Taxonomic metagenome profiling used ResPipe^12^ (see Methods), classifying a median of 72% (IQR:50-86) of sequencing reads as bacterial. Sink-ome communities were highly polymicrobial, with substantial variation across sink-traps (Extended data Fig.2). A total of 8845 unique bacterial species were identified across all sink-traps with a median estimated richness (Chao1) of 3520 species per sink (IQR:2324-4265). However, species composition varied substantially between sink-traps with respect to community diversity (Shannon index; median 4.38; IQR:3.48-5.50) and evenness (Pielou’s evenness; median 0.55; IQR:0.45-0.67), indicating heterogenous sink-omes varying in species content and abundance.

**Figure 2.**
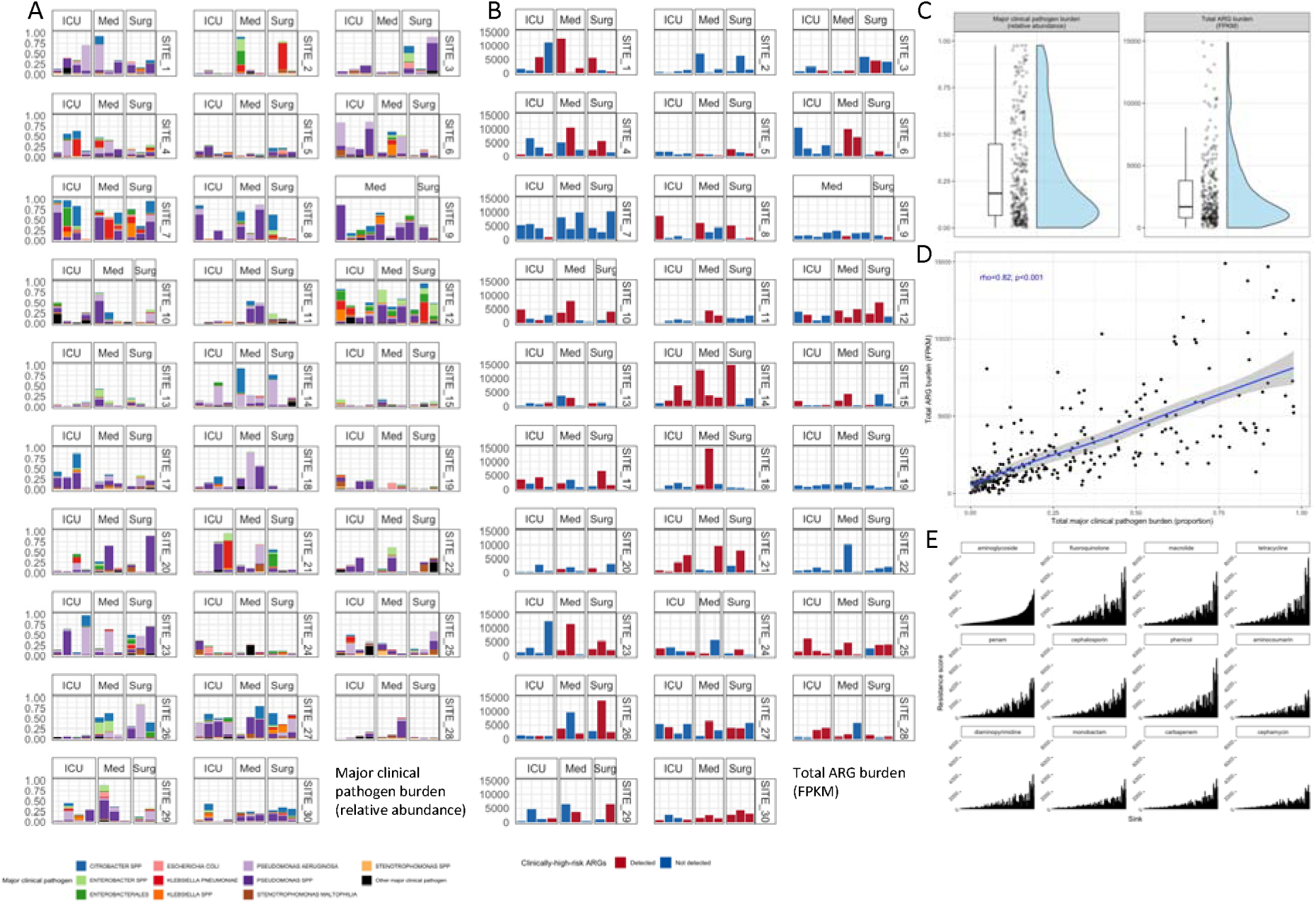
Distribution of major clinical pathogen and antimicrobial resistance gene (ARG) burden in sink-traps. A, Relative abundance (y-axis) of major clinical pathogen groups (colour) by individual sink-trap faceted by de-identified hospital site code and ward type. B, FPKM (fragments per kilobase million) (y-axis) of total ARG burden by individual sink-trap faceted by de-identified hospital site code and ward type, with colour indicating if clinically-high-risk ARGs were detected. C, Distribution of total major clinical pathogen burden and total ARG burden across all sink-traps. D, Relationship between total ARG burden (FPKM [y-axis]) and total relative abundance of major clinical pathogens (x-axis) by sink-trap (points), with Spearman’s rho and significance labelled and gam smooth trend line fitted with 95% confidence interval. E, Antibiotic class-level resistance (FPKM; y-axis) per sink-trap (x-axis) for the top 12 antibiotics (facets). The ordering of sink-traps along the x-axis is based on aminoglycoside resistance FPKM and then consistent across facets.

### Distribution of major clinical pathogen groups in sink-omes

To focus on clinically-relevant species, we considered a sub-group of 21 “major clinical pathogens” (see Methods) commonly linked to healthcare-associated and/or drug-resistant infections and including high-risk ESKAPEE pathogens^13^. All sink-traps contained ≥5 major clinical pathogens but the relative abundance of these varied substantially between sink-traps and across hospitals (median relative abundance of reads classified as major clinical pathogens: 0.18 [IQR:0.07-0.45, range:0.00042-0.98]). Some sink-traps were minimally colonised with, and some almost completely dominated by, major clinical pathogen groups (Fig.2A, 2C). Non-aeruginosa Pseudomonas spp. and Pseudomonas aeruginosa were identified in all sink-traps and were the most abundant group overall (Table 1, Extended Data Fig.3), with wide variation in observed abundances (median relative abundance: 0.037 [IQR:0.012-0.12] and 0.012 [0.0039-0.038]; range: 0.00001-0.87 and 0.00001-0.88, respectively). Similarly, Stenotrophomonas maltophilia and Escherichia coli were identified in all sink-traps, but at lower abundance (Table 1). Sink-trap colonisation with other healthcare-infection-associated Gram-negative bacilli was also common (92-99% sink-traps; Table 1).

**Table 1:** Relative abundance and prevalence (number [%] of sink-traps positive) for major clinical pathogen groups identified in sink-trap metagenomes. Groups are exclusive, i.e. “Enterobacterales” is all Enterobacterales other than those shown.

| Major clinical pathogen group | Relative abundance |  | Prevalence |
| --- | --- | --- | --- |
|  | Median (IQR) | Range | Sink-traps positive |
| *Pseudomonas* spp | 0.03700 (0.01200–0.12000) | 0.00001–0.86737 | 287/287 (100%) |
| *Pseudomonas aeruginosa* | 0.01200 (0.00390–0.03800) | 0.00001–0.88243 | 287/287 (100%) |
| *Stenotrophomonas maltophilia* | 0.00280 (0.00120–0.00850) | 0.00002–0.19630 | 287/287 (100%) |
| Enterobacterales | 0.00210 (0.00076–0.00640) | 0.00000–0.44401 | 285/287 (99%) |
| *Enterobacter* spp | 0.00200 (0.00055–0.01000) | 0.00000–0.31779 | 284/287 (99%) |
| *Stenotrophomonas* spp | 0.00170 (0.00065–0.00400) | 0.00000–0.10593 | 285/287 (99%) |
| *Citrobacter* spp | 0.00160 (0.00028–0.01500) | 0.00000–0.59865 | 284/287 (99%) |
| *Escherichia coli* | 0.00120 (0.00044–0.00400) | 0.00001–0.18002 | 287/287 (100%) |
| *Klebsiella pneumoniae* | 0.00100 (0.00036–0.00480) | 0.00000–0.66742 | 285/287 (99%) |
| *Acinetobacter* spp | 0.00034 (0.00005–0.00370) | 0.00000–0.21796 | 263/287 (92%) |
| *Klebsiella* spp | 0.00034 (0.00012–0.00170) | 0.00000–0.33008 | 281/287 (98%) |
| *Serratia marcescens* | 0.00012 (0.00005–0.00022) | 0.00000–0.25979 | 276/287 (96%) |
| *Acinetobacter baumannii* | 0.00008 (0.00003–0.00042) | 0.00000–0.05837 | 276/287 (96%) |
| *Enterococcus* spp | 0.00006 (0.00002–0.00019) | 0.00000–0.03284 | 78/287 (27%) |
| *Escherichia* spp | 0.00005 (0.00002–0.00028) | 0.00000–0.00268 | 196/287 (68%) |
| *Staphylococcus aureus* | 0.00003 (0.00001–0.00009) | 0.00000–0.00703 | 201/287 (70%) |
| *Enterococcus faecalis* | 0.00002 (0.00001–0.00012) | 0.00000–0.01334 | 71/287 (25%) |
| *Enterococcus faecium* | 0.00002 (0.00001–0.00013) | 0.00000–0.00671 | 72/287 (25%) |
| *Morganella morganii* | 0.00002 (0.00001–0.00004) | 0.00000–0.00087 | 230/287 (80%) |
| *Clostridioides difficile* | 0.00001 (0.00001–0.00002) | 0.00000–0.00022 | 69/287 (24%) |
| *Proteus* spp | 0.00001 (0.00001–0.00002) | 0.00000–0.00031 | 103/287 (36%) |

In contrast to the widespread presence of major pathogenic Gram-negative bacilli in sink-omes, Clostridioides difficile, Enterococcus faecalis, Enterococcus faecium and other Enterococcus spp. were identified in ∼25% of sink-traps. As a marker of recent faecal contamination we also screened for Crassvirales (human gut-associated phage), detected in 142/287 (49%) sinks (Extended Data Fig.3). Crassvirales abundance was positively associated with 10/21 major clinical pathogens, including S. aureus (Spearman’s rho=0.28 [95% CI:0.16-0.39], p<0.001), C. difficile (0.22 [0.09-0.33], p<0.001) and Proteus spp. (0.21 [0.09-0.32], p<0.001), possibly reflecting transient post-seeding events of sink-traps with some of these gut-associated species compared with persistent colonisation by others.

### Antimicrobial resistance gene burden in sink-traps

ARG profiling used the Resistance Gene Identifier (RGI-bwt) with the CARD protein homolog model; ARG counts were normalised by calculating Fragments Per Kilobase Million (FPKM) (see Methods). We then calculated total ARG burden (FPKM sum of all ARGs classified) and a separate “clinically high-risk ARG” burden (FPKM sum of all bla_CTX-M_ genes [major extended-spectrum beta-lactamases] plus the major five carbapenemase families: bla_IMP_, bla_KPC_, bla_OXA-48_-like genes, bla_NDM_, bla_VIM_). A total of 2392 unique ARGs were detected across sink-traps, comprising 286 ARG families. The median estimated ARG richness (Chao1) per sink-trap was 170 ARGs (IQR:124-215) (Extended Data Fig.2). However, overall sink-trap ARG composition was less varied than species composition (Shannon diversity: median 3.88; IQR:3.49-4.17; Pielou’s evenness: median 0.77; IQR:0.73-0.80). Similar to major clinical pathogen burden, total ARG burden varied considerably across individual sink-traps (Fig.2B, 2C), with a median of 1677 FPKM (IQR:805-3793) and wide variation across sink-traps (range:0-14879 FPKM).

Clinically-high-risk ARGs were detected in 115/287 (40%) of sink-traps (median FPKM:3.03 [IQR:0.48-32.7]) and across 26/29 (90%) of hospitals (hospital-level median FPKM:66 [IQR:38-150]). bla_CTX-M_ was detected in 56/287 [20%] of sink-traps and major carbapenemases in 5.2-11% of sink-traps (Table 2). bla_KPC_ was the most abundant clinically high-risk ARG (median FPKM: 25 [IQR:4.6-39]; Table 2).

**Table 2.** Fragments per Kilobase Million (FPKM) and prevalence of clinically-high-risk ARGs across sink-traps and hospitals.

| Clinically-high-risk ARG | FPKM | Prevalence |  |
| --- | --- | --- | --- |
|  | Median (IQR) | Sink-traps positive | Hospitals positive |
| KPC carbapenemase | 25.00 (4.60–39.00) | 17/287 (5.9%) | 11/29 (38%) |
| OXA-48-like carbapenemase | 4.40 (1.20–18.00) | 26/287 (9.1%) | 14/29 (48%) |
| VIM carbapenemase | 3.80 (0.86–36.00) | 33/287 (11%) | 14/29 (48%) |
| CTX-M beta-lactamase | 1.30 (0.30–6.00) | 56/287 (20%) | 21/29 (72%) |
| IMP carbapenemase | 0.65 (0.10–1.40) | 25/287 (8.7%) | 17/29 (59%) |
| NDM carbapenemase | 0.26 (0.10–1.40) | 15/287 (5.2%) | 11/29 (38%) |

When stratifying by antibiotic class-level resistance (see Methods), ARGs conferring resistance to 12 clinically-relevant antibiotic classes were detected in almost all sink-traps (96-99%) (Extended Data Table 1). ARGs encoding aminoglycoside and fluroquinolone resistance were most abundant (median FPKM: 556 [IQR:257-1030] and 541 [212-1322], respectively). Like major clinical pathogen abundance, antibiotic class-level resistance varied widely by sink-trap. However, sink-traps with higher FPKM of ARGs conferring resistance to one antibiotic class typically exhibited higher FPKMs of ARGs conferring resistance to multiple antibiotic classes, consistent with co-selection (Fig.2E). All 12 antibiotic class-level resistance FPKMs were significantly co-correlated at the sink-trap-level (Spearman’s Rho median: 0.93; IQR:0.90-0.96; all p-value<0.001) (Extended Data Fig.4). This contrasted with major clinical pathogen groups (excluding correlations within-genus/order) where a high-burden single clinical pathogen tended to dominate the sink-trap, consistent with intra-niche competition rather than co-selection (Spearman’s Rho median: 0.15 [IQR:0.06-0.62]; p-value median: 0.009 [IQR:<0.001-0.30]; (Extended Data Fig.5).

### Associations between sink-trap taxonomic and resistome composition

Total ARG burden in sink-traps was significantly associated with total major clinical pathogen burden (Spearman’s Rho:0.82 [95% CI:0.77-0.86]; p<0.001), with sink-traps with a low pathogen burden having lower ARG burdens (Fig.2D), consistent with ARGs being concentrated in clinical pathogens. For clinically-high-risk ARG burden, the association was similar but weaker (Spearman’s Rho:0.22 [0.11-0.32]; p<0.001). As the total major clinical pathogen burden increased across sink-traps, overall bacterial species richness (Chao1: Spearman’s Rho:-0.35 [-0.45,-0.23]; p<0.001) and diversity (Shannon) declined (−0.52 [-0.62,-0.41]; p<0.001) (Extended Data Fig.6). Bacterial species diversity and richness also declined (Shannon:-0.55 [-0.63,-0.44]; p<0.001; Chao1:-0.34 [-0.46,-0.23]; p<0.001) with increasing total ARG FPKM, again consistent with the strong positive correlation observed between major clinical pathogen and ARG burden (Fig.2C).

Non-metric Multidimensional Scaling (NMDS) showed limited hospital-level clustering (Extended Data Figs.7, 8), reflecting that some hospitals were consistently high in major clinical pathogen burden across all sinks (e.g. sites 7, 12) and others consistently low (e.g. site 15) (Figs.2A, 2B). However, there was no clear clustering of sink-ome profiles by ward or sink location for either bacterial species or ARG composition, indicating sink-trap and hospital-level factors were likely driving variation. NMDS for sink-trap bacterial species and resistome were significantly positively associated (PROTEST: p<0.001; r=0.60), again consistent with the association between pathogen and ARG burdens (Fig.2D).

### Association between sink-trap sampling, setting and features, and bacterial species and resistome composition

To evaluate associations between 49 sink-associated features (including six core design features and 43 other explanatory variables; see Supplementary dataset) and major clinical pathogen/ARG burden (Extended Data Fig.9), we fitted Generalised Additive Models (GAMs) with AIC-based backwards elimination and site-level random effects (see Methods, Supplementary dataset). We focused our interpretation of results on variables with very high supporting evidence of effect (p<0.001). If a variable met this threshold in any model, we also considered its effects with p<0.05 in other models (model estimates and confidence intervals available in the Supplementary dataset).

1. Impact of sink-trap antibiotics and chemistry on sink-omes

Detection of beta-lactam antibiotics was strongly associated with a higher major clinical pathogen burden in sink-omes (relative abundance:+0.51 [95% CI:+0.24,+0.78], p<0.001), total ARG burden (log_10_FPKM:+0.12 [+0.02,+0.22], p=0.02) and clinically-high-risk ARG burden (FPKM:+0.67 [+0.05,+1.29], p=0.03) (Fig.3). Similarly, iron readings of 5mg/L (versus 0) were associated with a higher clinically-high-risk ARG burden (FPKM:+1.78 [+0.75,+2.80], p<0.001) and higher major clinical pathogen burden (relative abundance:+0.53 [+0.07,+0.99], p=0.02). The highest sink-trap alkalinity readings of 240mg/L were strongly associated with higher clinically-high-risk ARG burden (FPKM:+5.30 [+2.59,+8.00], p<0.001), with less strong associations for 180mg/L (FPKM:+3.56 [+1.31,+5.80], p=0.002), 80mg/L (FPKM:+1.68 [+0.10,+3.26], p=0.03) and 40mg/L (FPKM:+1.94 [+0.66,+3.23], p=0.003), suggesting a concentration-dependent relationship.

ii) Impact of sink sample characteristics on sink-omes

**Figure 3.**
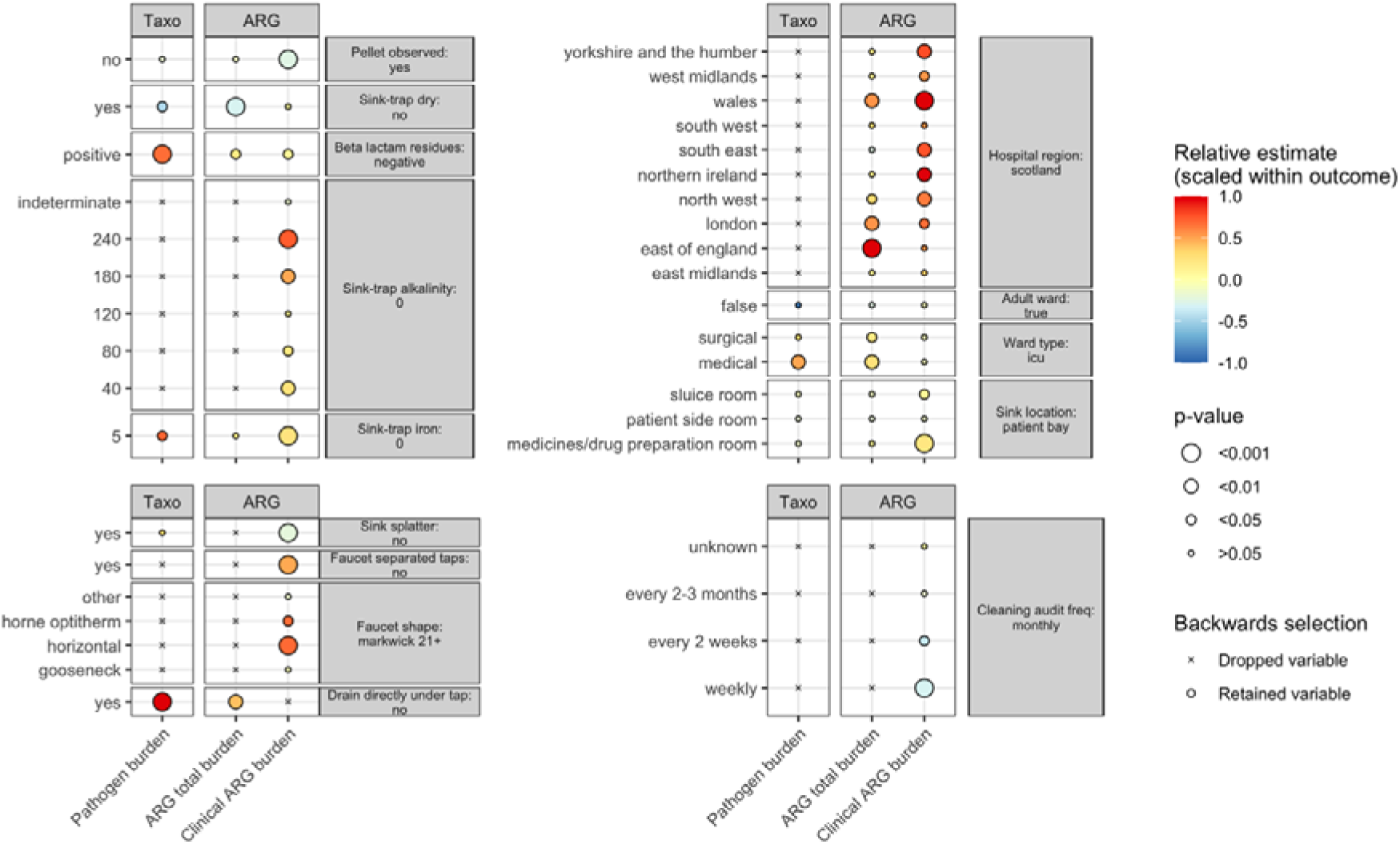
Associations between sink features and major clinical pathogen/antimicrobial resistance gene (ARG) burden. Estimates from backward selection for the three major outcomes considered (x axis/facets); plot limited to variables with at least one estimate with very strong support (p<0.001) or considered core study design features (see supplementary material for full model estimates). Reference levels are indicated in right facets with comparator levels labelled on left. Shape reflects whether the variable was eliminated during backwards selection, size of circle reflects the significance of estimate and colour indicates relative effect size.

Sink-ome composition was influenced by whether traps were water-filled or dry during the initial aspiration attempt (i.e. requiring refilling with tap water pre-sampling). Compared to water-filled sink-traps, bacterial communities in dry sink-traps had lower total ARG burden (log_10_FPKM:-0.18 [-0.29,-0.08], p<0.001) and lower major clinical pathogen (relative abundance:-0.35 [-0.65,-0.06], p=0.01). Additionally, sink-trap aspirates which did not produce a pellet post-centrifugation (consistent with less planktonic bacteria) had lower clinically-high-risk ARG burden (FPKM:-1.85 [-2.70,-1.00], p<0.001).

iii) Impact of reported sink cleaning and usage on sink-omes

Cleaning audit frequency was associated with variation in both species and ARG composition of sink-traps. Most sinks were cleaned with chlorine-based products (216/287 [75%]); citric acid-based, alcohol alkoxylate-based, and unknown cleaning product use was reported for 56 (20%), 12 (4%) and 23 (8%) sinks, respectively. Compared with monthly audits, audits conducted weekly or fortnightly were linked with lower clinically-high-risk ARG burden (FPKM:-2.30 [-3.55,-1.04], p<0.001; −2.71 [-5.16,-0.26], p=0.03), respectively.

iv) Impact of sink design features on sink-omes

Sinks designed such that tap water flowed directly onto drains were associated with higher major clinical pathogen burden (relative abundance:+0.76 [+0.33,+1.18], p<0.001) and total ARG burden in sink-omes (log_10_FPKM:+0.24 [+0.08,+0.40], p=0.003). Specific sink faucet designs were also linked to higher clinically-high-risk ARG burden in sink-traps, namely separated hot/cold taps (FPKM:+3.52 [+1.99,+5.10], p<0.001), horizontal-style faucets (versus Markwik 21+) (FPKM:+5.00 [+2.23,+7.76], p<0.001) and Horne Optitherm (FPKM:+4.91 [+0.46,+9.35], p=0.03). We also found lower major clinical ARG burden (FPKM:-1.57 [-2.48,-0.67], p<0.001) in sinks with evidence of sink splatter.

ii) Impact of sink-trap location within hospitals on sink-omes

Sink-traps located in medicines/drug preparation rooms (versus patient bays) were associated with a higher burden of clinically-high-risk ARGs (FPKM:+1.6 [+0.74,+2.45], p<0.001). Sluice room sink-traps were also associated with higher clinically-high-risk ARG burdens (FPKM:+0.86 [+0.05,+1.68], p=0.04). For geographic region, higher ARG total burden was associated with the East of England (log_10_FPKM:+0.59 [+0.32,+0.87], p<0.001), London (+0.33 [+0.09,+0.57], p=0.008), Wales (+0.33 [+0.13,+0.53], p=0.002) and the North-west (+0.19 [+0.02,+0.35], p=0.027) (versus Scotland). Higher clinically-high-risk ARG burdens were associated with hospital sink-traps in Wales (FPKM:+7.28 [+3.65,+10.92], p<0.001), Yorkshire and the Humber (+5.68 [+2.03,+9.34], p=0.003), West Midlands (+3.98 [+0.47,+7.49], p=0.027), South East (+5.53 [+1.51,+9.54], p=0.008), Northern Ireland (+7.20 [+2.31,+12.09], p=0.004), North West (+4.58 [+1.60,+7.56], p=0.003), and London (+4.90, [+0.03,+9.76], p=0.05).

## Discussion

Our evaluation of 287 sink-traps across 29 UK hospitals reveals that important pathogenic species and ARGs are near-ubiquitous in UK hospital sink-traps, with substantial heterogeneity in quantitative burden. Some sink-omes consist almost entirely of major clinical pathogens, and notably, high pathogen burdens correlate with high ARG burdens. 40% of sink-traps harboured either CTX-M type ESBLs or major carbapenemases. The variability in pathogen and ARG sink-trap burden was driven to some degree by hospital, potentially related to differences in patient populations and hospital-level factors. However, most of the variability was driven by specific sink/sink-trap-related factors, similar to findings in a previous study considering isolate-level genomic variation in Enterobacterales in patients and hospital environmental reservoirs^14^. Exploring this further, we identified significant associations between sink-trap pathogen/ARG burden and sink-level features such as sink design, location, sink-trap chemistry and cleaning protocols, which likely influence biofilm formation, and pathogen and ARG selection and persistence. A key question is whether these factors could represent targets for interventions to mitigate the pathogen/ARG burden observed in sink-traps and thereby the risk to patients, although importantly our results reflect associations that are not necessarily causal.

The detection of at least five major clinical pathogens in sampled sink-traps strongly supports the known role of the hospital wastewater environment as a pathogen reservoir contributing to patient colonisation and infection^5^. Notably, Pseudomonas aeruginosa, Stenotrophomonas spp. and clinically-relevant Enterobacterales were found in almost all (98-100%) sinks and are common healthcare infection-associated pathogens. Pseudomonas spp. and Stenotrophomonas were frequently the most abundant species in sink-omes, in keeping with at least one other smaller study^15^, likely driven by the ability of these bacteria to form resilient biofilms in moist environments^16^. Studies focused on sequencing P. aeruginosa and drug-resistant Enterobacterales isolates to understand transmission between patients and sinks have estimated sink-to-patient acquisition accounts for 7-40% of transmissions^14,17,18^ (although these were mostly asymptomatic gastrointestinal colonisation), with patient-to-sink dissemination accounting for 45% of cases^14^. The latter finding would be consistent with Crassvirales (a human faeces-specific marker) detection in ∼50% of sinks in this study.

Like the distribution of major clinical pathogens, nearly all sinks harboured ARGs encoding resistance to the 12 antibiotic classes considered, with strong positive associations in cross-class ARG abundance, albeit with highly variable total ARG abundance observed sink-to-sink. Interestingly, ARGs co-occurred irrespective of the dominance of major clinical pathogens in some sink-omes, suggesting they are maintained together regardless of the species-level competition shaping these communities, and consistent with the fact that many may be horizontally mobilised on mobile genetic elements. Our finding of significant associations between reduced microbiome diversity and increased ARG and major clinical pathogen burdens suggests that sink-ome dysbiosis may be both a consequence and driver of colonisation by ARG-associated clinical pathogens. These dynamics closely resemble the disruption of colonisation resistance in human gut microbiomes which can promote infections such as C. difficile and colonisation with drug-resistant Enterobacterales^19,20^, and where recurrence has been effectively treated with attempts to restore a “healthy microbiome” through biotherapeutic products and microbiota transplantations^21^. Approaches aimed at positively modulating the sink-trap microbiome could serve as effective interventions for both preventing and mitigating sink-trap colonisation with antimicrobial-resistant and/or healthcare associated pathogens.

Based on our observations, sink-trap chemistry likely plays a major role in shaping pathogen and ARG burdens in sink-trap microbiomes and could represent an opportunity for intervention. Sinks positive for beta-lactam residues exhibited increased major clinical pathogen, total ARG burdens and high-risk clinical ARG burden, and likely represent a major selection pressure for drug-resistant pathogens in these reservoirs. In this study, 95/287 (33%) sink trap aspirates had at least one antibiotic class detected, similar to another study^22^, and 87/287 (30%) of sink-traps contained detectable beta-lactams^23^. Strategies to ensure that medicines (including antibiotics) and patient waste containing antimicrobial metabolites are not disposed of via sinks should be prioritised for implementation.

Iron and alkalinity were also associated with sink-trap microbial community structure. Sink-traps with iron measurements at 5mg/L (versus 0) had higher major clinical pathogen, total ARG, and clinically-high-risk ARG burdens. Elevated iron levels can facilitate bacterial growth and biofilm formation^24^. Iron/steel pipes in premise plumbing can leach iron to water in a range of 0.2–18 mg/L^25^; given the long timeframe over which NHS infrastructures have been developed it is plausible that some pipework in UK hospitals contains iron. This could also represent a consideration for any future infrastructure projects. We also observed a concentration-dependent effect for alkalinity whereby high alkalinity was independently associated with higher burdens of clinically-high-risk ARGs. High alkalinity may reduce the effectiveness of cleaning approaches which are often pH-dependent such as chlorine-releasing agents (used in 75% of sinks in this study)^26^.

Samples obtained from “dry” sink traps and without a visible pellet post-centrifugation were associated with lower pathogen, total ARG and clinically-high-risk ARG burdens; thus keeping sink-traps dry through improved drainage^27^ or heating may represent useful interventions, consistent with studies showing reductions in Enterobacterales and P. aeruginosa colonisation following heat-based sink-trap interventions^28–30^.

Cleaning audits conducted more frequently than monthly were associated with lower clinically-high-risk ARG burdens suggesting that structured cleaning regimes including frequent audits may be beneficial for mitigating sink-trap colonisation in addition to reducing surface contamination. However, we also found some evidence (p<0.01) that intensively cleaned (>1xday) sink-traps had an increased major clinical pathogen compared with those cleaned daily (Supplementary material), suggesting that excessive cleaning may paradoxically contribute to pathogen and AMR selection; this has also been shown in recent experimental studies^31–34^. Capturing more detailed and informative data on specific cleaning regimes and products used was not possible in this study, but is likely to be relevant.

Several sink design features were associated with differential impacts on pathogen and ARG burden. Although these are unlikely to be directly influencing microbial communities, they may be reflecting unmeasured drivers, such as water flow (faucet shape) and temperature (separated hot/cold taps), or other unmeasured aspects of usage. For instance, tap streams directly entering drains may result in aeration of sink-trap wastewater or biofilm disruption contributing to higher major clinical pathogen burdens. Interdisciplinary innovation with engineering teams and physicists would be helpful to optimise infrastructure design to minimise biofilm formation, pathogen overgrowth and dissemination risks from sinks^35,36^.

Sink-traps in staff-only medicine/drug preparation rooms exhibited high levels of clinically-high-risk ARGs. This potentially reflects medicine disposal into these sinks^37^ selecting for antibiotic-resistant organisms^11^, and consistent with the observation that medicine/drug preparation room sink-traps were most likely to be positive for antibiotics^23^. Although patients do not have direct contact with sinks in sluice or medicine/drug preparation rooms, the potential for sink-trap biofilms to spread throughout wards via interconnected plumbing^36^ and the risk of sink splatter contaminating staff or equipment^38^ suggests that these sinks may still pose a transmission risk to patients. Regional differences in sink-trap total ARG burden and/or clinically high-risk ARG burden were apparent, with sink-traps in several regions in England, and sink-traps in Wales and Northern Ireland, exhibiting significantly higher ARG burdens compared to those in Scotland. For the six affected regions in England (the North-West, East of England, Yorkshire and the Humber, West Midlands, South-East, and London regions), these were the same regions with the highest AMR rates observed during national bloodstream infection surveillance in 2023-2024^2^. Although most research on sink-trap environmental reservoirs has occurred in critical care settings and for antibiotic-resistant bacteria, the burden and contribution to transmission may be greater in non-critical care wards and relevant for susceptible healthcare-associated pathogens.

Our study has several limitations. It was undertaken during a single time window, with each sink-trap evaluated only at a single timepoint. Other studies have demonstrated that microbial composition can vary markedly over time, and that sinks within wards have more similar taxonomic profiles than those between wards suggesting distinct ward-based wastewater ecologies^6^. Our breadth of sampling across multiple hospitals UK-wide was a trade-off for investigating ward-level sink diversity in more limited detail. Sink-trap pathogen and ARG epidemiology may be different in other settings, as these are influenced by sink-trap specific factors. There are many different approaches to taxonomic and ARG profiling in metagenomes and it remains unclear which is the most accurate approach; however, importantly we applied a consistent approach within-study enabling relevant comparison. We were unable to evaluate the relationship between pathogen/ARG abundance in sink-trap microbiomes and patient colonisation burden or clinical infection-associated data in populations co-located with the sampled sinks, and the threshold at which sink-drain colonisation becomes a risk to patients remains unexplored - this is important future work. Similarly, we were unable to assess whether ward-level wastewater could be used to monitor pathogen/AMR burden in ward inpatients.

Overall, however, our findings have significant implications for optimising the design, cleaning, usage and monitoring of hospital sinks to ensure that selection pressures for pathogens and ARGs can be avoided, minimising the risk to patients. Our findings are also relevant to sinks in community settings where pathogen/ARG burdens may be even higher^39^, potentially contributing to community-acquired, hospital-onset infections. Our findings of sink-trap microbiome dysbiosis and associations with pathogen/ARG abundance warrant future research into what sink-ome features contribute to colonisation resistance against drug-resistant pathogens and how this might inform interventions. In the absence of removing sinks from healthcare completely, interventional studies to characterise the best strategies to manage them and minimise any negative impact on patients are urgently needed.

## Methods

### Sink survey design and data collection

For our survey, we partnered with the National Infection Team Collaborative for Audit and Research (NITCAR; https://nitcollaborative.org.uk/wp/), a UK-wide network of infection-focused healthcare professionals, to recruit 29 hospitals (Fig.1). Sites were instructed to collect data and sink-trap wastewater samples at one timepoint between Jan-Mar 2023 from 10 sinks in total, including four sinks in general critical care settings, three in general medical wards, and three in general surgical wards. The rationale was to consider setting-specific variability in common hospital settings, including differences in antibiotic prescribing, patient turnover, and patient-staff ratios. For each sink sampled, a standard set of metadata was collected through REDCap (https://www.project-redcap.org) including ward type, sink location within the ward (i.e. patient bay, patient side-room, sluice room, medicines/drug preparation room), distance of the sink to the nearest patient bed, sink usage type, evidence of splatter around the sink if the taps were turned on at a handwashing flow rate, sink cleaning frequency and cleaning products used. Each sink was also photographed to facilitate characterisation of specific sink design features based on guidance and published evidence suggesting that minimising biofilm formation, contamination, and splatter risk to patients and items would be beneficial, including: sink basin material type, sink basin shape, presence of plug, overflow and drain strainer, location of drain with respect to tap outlet, presence of a basin fin; location of taps, method of tap operation, and shape of the outlet spout. These features were classified independently by two researchers with disagreements resolved by consensus.

### Sink-trap aspirate sampling, chemical and antibiotic residue testing

Sink-trap aspirates were collected, and dipsticks were tested according to a standardised protocol) and instructional training video shared with site investigators (https://figshare.com/articles/media/SinkBug_-_instructional_training_video/29617739?file=56454293). In brief, sink-traps were aspirated using sterile 50ml syringes and nasogastric tubing before antibiotic/chemical dipstick testing at site. Aspirates were tested semi-quantitatively for beta-lactam, tetracycline, sulfonamide and quinolone antibiotic classes using the previously validated^40^ QuaTest BTSQ 4-in-1 rapid test kit (Ringbio, UK). Aspirates also underwent chemical testing for alkalinity, water hardness, and pH, and semi-quantitative measurement of chlorine, copper, fluoride, iron, lead, nitrate, nitrite (all mg/L) using the Bebapanda Upgrade 14-in-1 dipstick and silver concentrations (mg/L) using Sensafe Boris’s Silvercheck strips, also previously validated^40^. After dipstick testing, aspirates were mixed with boric acid to preserve bacterial community composition^40^, and shipped to Oxford for processing for metagenomic sequencing.

### DNA extraction, metagenomic sequencing and sequence data processing

Metagenomic DNA was extracted from sink-trap aspirates using the DNeasy PowerSoil kit (Qiagen) according to the manufacturer’s instructions. Library preparation and sequencing (paired-end 150bp) was conducted by the Oxford Genomics Centre using the NovaSeq6000 (Illumina) platform, to generate an expected yield of 10-20 million paired-end reads per sample (∼3.9 gigabases). This depth was chosen from randomly subsampling 40 million reads from 17 previously sequenced metagenomes from UK clinical handwash basin sinks to depths of 1 million, 2 million, 4 million, 6 million, 10 million and 20 million reads and considering AMR gene recovery. Rarefaction analyses demonstrated 10-20 million paired-end reads captured most AMR gene diversity (Extended Data Fig.9) reflecting a suitable trade-off between resource and sequencing depth. Raw metagenomic reads underwent QC, adapter trimming and filtering using Trim Galore^41^ (v0.6.10) with Phred-score and read length thresholds of 25 and 75bp respectively.

Taxonomic profiling used the taxonomic module of ResPipe^12^ (v1.6.1) which incorporates Kraken2^42^ and Bracken^43^ (Standard index v10/9/2023). We used Bracken-based re-estimation of abundance normalised to relative abundance in our taxonomic evaluations with the default reporting threshold of 0.001% relative abundance. To focus on clinically-relevant species, we considered a sub-group of “major clinical pathogens” chosen based on clinical expertise and reflecting bacterial pathogens included in the national English microbiology linked dataset, the Second Generation Surveillance System (SGSS), namely: Acinetobacter baumannii, non-baumannii Acinetobacter spp., Citrobacter spp., Clostridioides difficile, Enterobacter spp., Enterococcus faecalis, Enterococcus faecium, non-faecalis and non-faecium Enterococcus spp., Escherichia coli, non-coli Escherichia spp., Klebsiella pneumoniae, non-pneumoniae Klebsiella spp., Morganella morganii, Proteus spp., Pseudomonas aeruginosa, non-aeruginosa Pseudomonas spp., Serratia marcescens, Staphylococcus aureus, Stenotrophomonas maltophilia and non-maltophilia Stenotrophomonas spp. Less common Enterobacterales genera included within SGSS were aggregated under other Enterobacterales (Raoultella, Kosakonia, Phytobacter, Leclercia, Salmonella, Kluyvera, Cronobacter, Lelliottia, Pluralibacter, Yokenella, Shigella, Serratia, Yersinia, Rahnella, Ewingella, Pantoea, Erwinia, Tatumella, Pectobacterium, Providencia, Moellerella, Edwardsiella, Hafnia, Obesumbacterium, Leminorella). All groups were exclusive (e.g. abundance of non-E. coli Escherichia spp. did not include Escherichia coli). This set of major clinical pathogens considered also included the ESKAPEE group (Enterococcus faecium, Staphylococcus aureus, Klebsiella pneumoniae, Acinetobacter baumannii, Pseudomonas aeruginosa, Enterobacter spp., and Escherichia coli).

ARG profiling used Resistance Gene Identifier (RGI)^44^ in metagenomic read mode (BWT) with the CARD protein homolog model (v3.3.0). ARG counts were normalised to sequencing depth and mapping target reference lengths by calculating Fragments Per Kilobase Million (FPKM); briefly, the depth-normalised abundance of each ARG was multiplied by 1,000,000 (Fragment/Reads Per Million) before division by the relevant ARG reference length in kb. We considered the total ARG burden (FPKM) by summing the FPKM for all known ARGs classified. We also considered AMR burden at the antibiotic class-level, by aggregating ARGs into categories associated with resistance to specific antibiotic classes using mappings in the CARD database (“Drug class” categories) which are based on experimental evidence of elevated minimum inhibitory concentrations (MICs) in the presence of specific mechanisms: namely to aminocoumarins, aminoglycosides, carbapenems, cephalosporins, cephamycins, diaminopyrimidines, fluoroquinolones, macrolides, monobactams, penams, phenicols and tetracyclines. Thirdly, we considered specific “major clinical ARGs” as a separate category by calculating the sum of FPKM over all bla_CTX-M_genes (the most clinically relevant extended-spectrum beta-lactamase group conferring resistance to third generation cephalosporins) and the major five carbapenemase families: bla_IMP_, bla_KPC_, bla_OXA-48-like_ genes, bla_NDM_, bla_VIM_.

### Statistical methods Exploratory ordination

We used Non-metric Multidimensional Scaling (NMDS) to explore sink-trap taxonomic and resistome composition at the species- and ARG-levels. Bray-Curtis dissimilarities were used as the distance metric for ordination, with 200 permutations. Stress plots were inspected to confirm that a non-metric fit was optimal. Procrustes analysis and PROTEST were used to assess and test the significance of concordance between taxonomic and resistome NMDS solutions.

Generalised Additive Modelling to assess metagenomic associations with sink features We considered three outcomes, namely a taxonomic outcome of major clinical pathogen burden (relative abundance), and ARG outcomes of total ARG burden (log_10_FPKM) and clinically-high-risk ARG burden (FPKM). All outcomes were truncated at the 5^th^ and 95^th^ percentile to reduce the impact of outliers and overfitting (Extended Data Fig.10).

To assess the association between each taxonomic/ARG outcome and sink features, we estimated Generalised Additive Models (GAMs) controlling for pre-specified study design features (ward type, adult/paediatric ward, sink location) and sampling characteristics (sink-trap dryness, observable pellet, sample volume), plus a random effect for hospital site. We used beta GAMs to evaluate associations with major clinical pathogen burden (bounded by 0-1); Gaussian GAMs to evaluate associations with total ARG burden (log10-transformed); and Tweedie GAMS to evaluate the association with clinically-high-risk ARG burden (due to zero-inflation). For each categorical variable, the reference level was defined as the level with the most observations. We then used backwards elimination based on the Akaike Information Criterion (AIC) to select a final model from all non-core variables, retaining design feature and sampling characteristic variables regardless of significance.

Extended data – see separate extended data document.

Supplementary tables – see separate supplementary document.

## Data availability

Raw sequencing data are available under NCBI BioProject PRJNA1190863, and sample accessions listed in Supplementary tables.

## Code availability

All analyses and plots may be reproduced using code and processed data available via GitHub at https://github.com/KaibondChau/Chau_etal_SinkBug where a knitted markdown can also be viewed.

## Ethics

With reference to the UK Health Research Authority’s algorithm, available at http://www.hra-decisiontools.org.uk/research/ and attendant leaflet, Defining Research, or by reference to The Health Care Quality Improvement Partnership (HQIP)’s Guide for Clinical Audit, or by reference to The Health Care Quality Improvement Partnership (HQIP)’s Guide for Clinical Audit, Research and Service review, it was determined that although our study was research, given that it involved no human participants, human tissue or personal data, it was not subject to the Department of Health’s UK Policy Framework for Health and Social Care Research (2017). It therefore was not deemed to require sponsorship or research ethics review.

## Data Availability

All data produced are available online at the relevant links (please see Data Availability links)

https://figshare.com/articles/media/SinkBug_-_instructional_training_video/29617739?file=56454293

https://www.ncbi.nlm.nih.gov/bioproject/?term=PRJNA1190863

https://github.com/KaibondChau/Chau_etal_SinkBug

## Acknowledgements

We are grateful to all the participating hospitals and the NITCAR committee for enabling the study, and to K. Arunachalam, N. Forbes, S. Fudge, L. Gargee, M. Kyffin, A. Maxwell, H. Reddy, and J. Vasant. We also wish to make a special acknowledgement of the work and contribution of Dr A. Bali.

This study was funded by the National Institute for Health Research (NIHR) Health Protection Research Unit in Healthcare Associated Infections and Antimicrobial Resistance (NIHR200915), a partnership between the UK Health Security Agency (UKHSA) and the University of Oxford, and was supported by the NIHR Oxford Biomedical Research Centre (BRC). The views expressed are those of the authors and not necessarily those of the NIHR, UKHSA or the Department of Health and Social Care. Christopher Darlow is funded by an NIHR Award (CL-2022-07-001).

## Author information

The SinkBug Consortium is represented by:

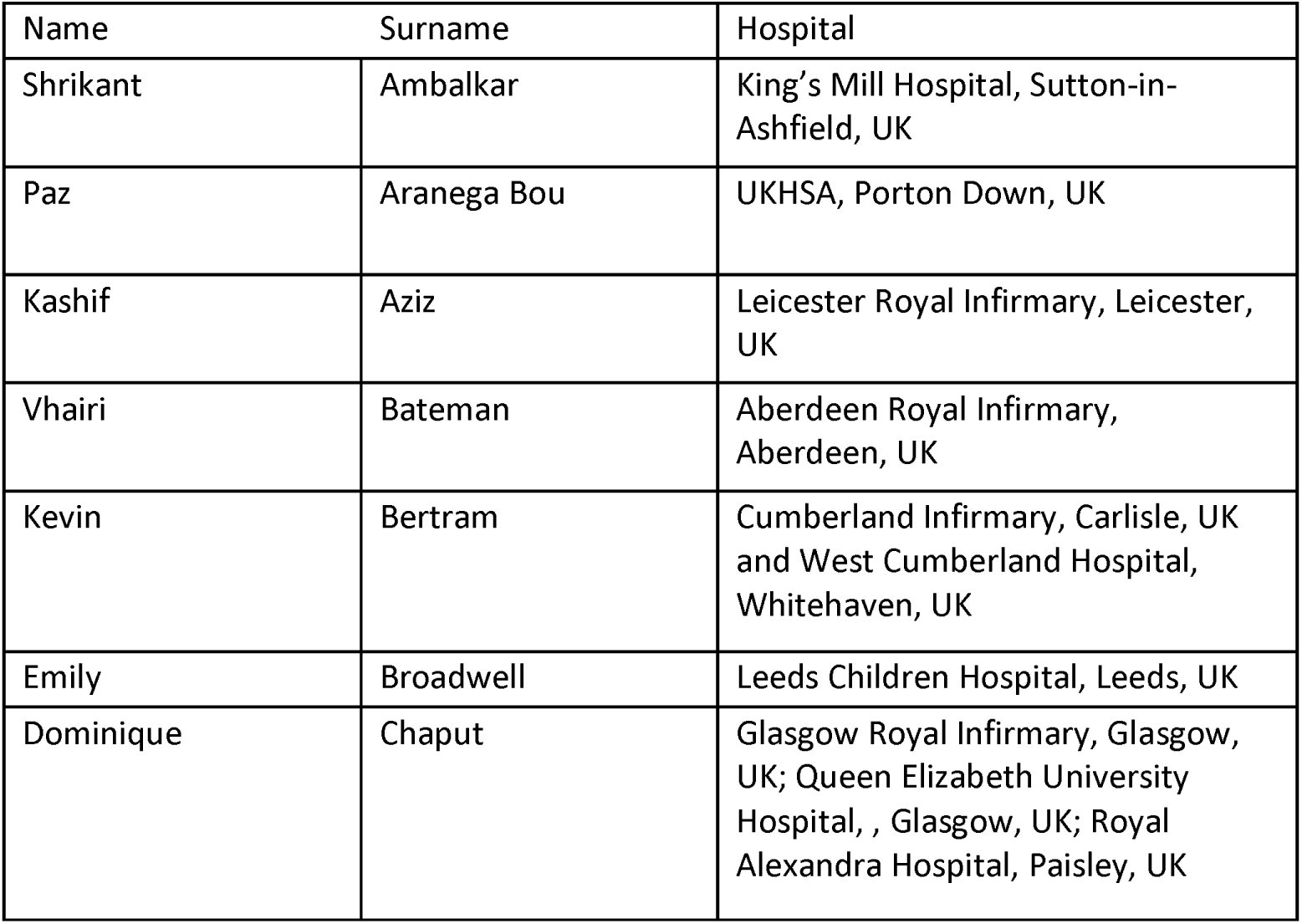
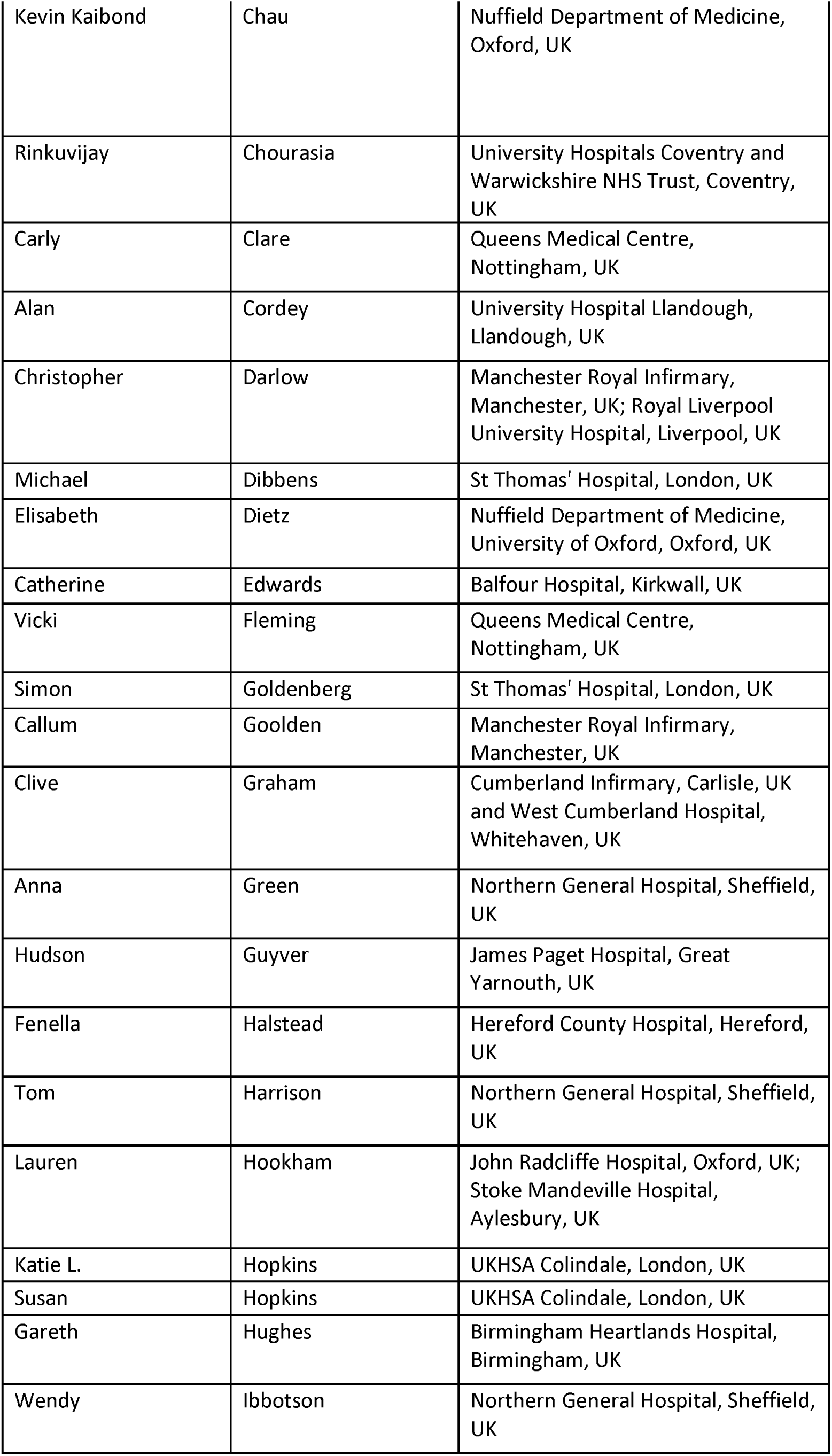
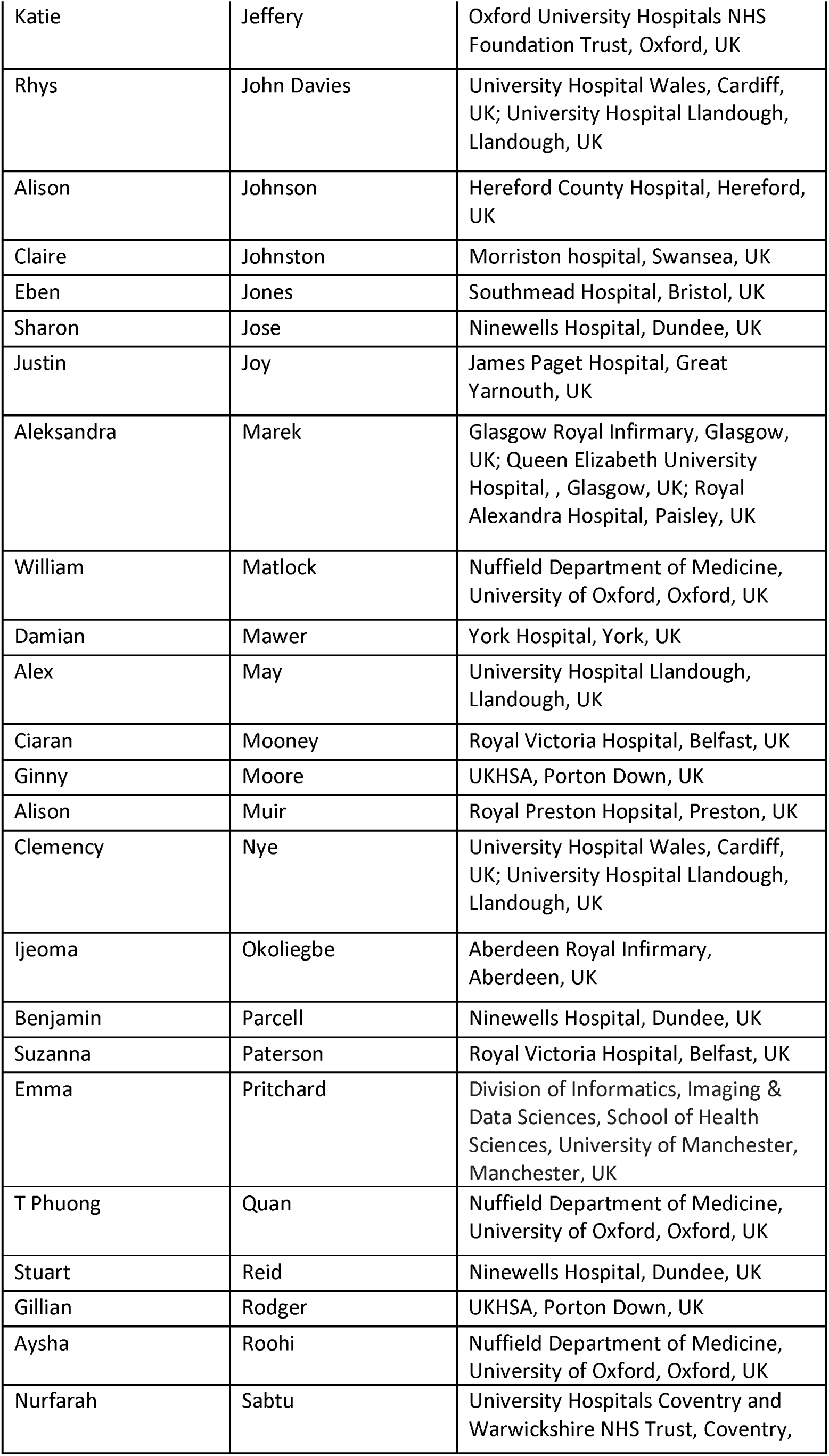
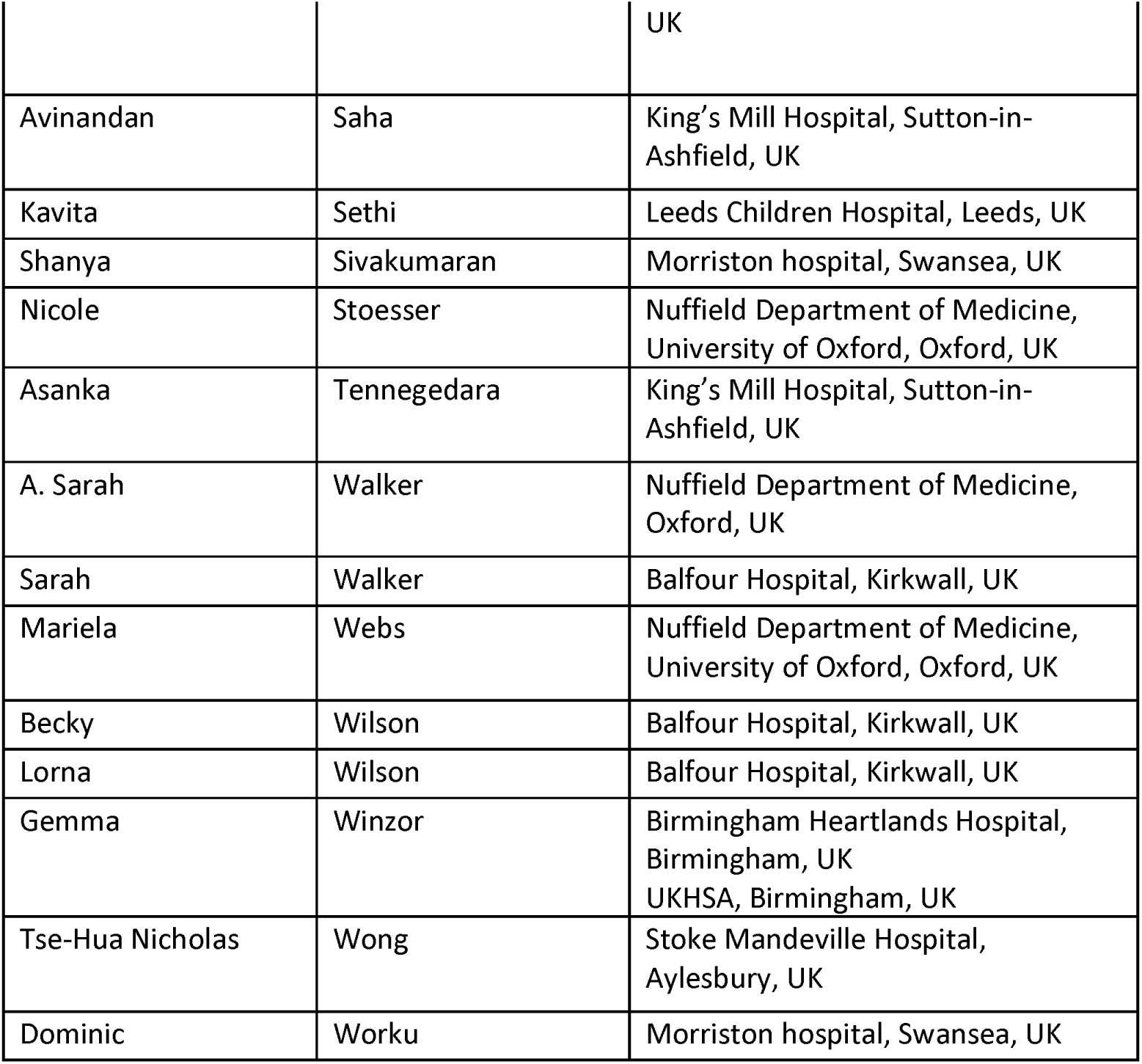

The authors have no conflicts of interest.

