## Extended data for "The hospital “Sink-ome”: Pathogen and antimicrobial resistance gene burden in sink-traps across 29 UK hospitals and associations with sink characteristics"

**Extended data items (Extended data Figures 1-10 and Table S1)**

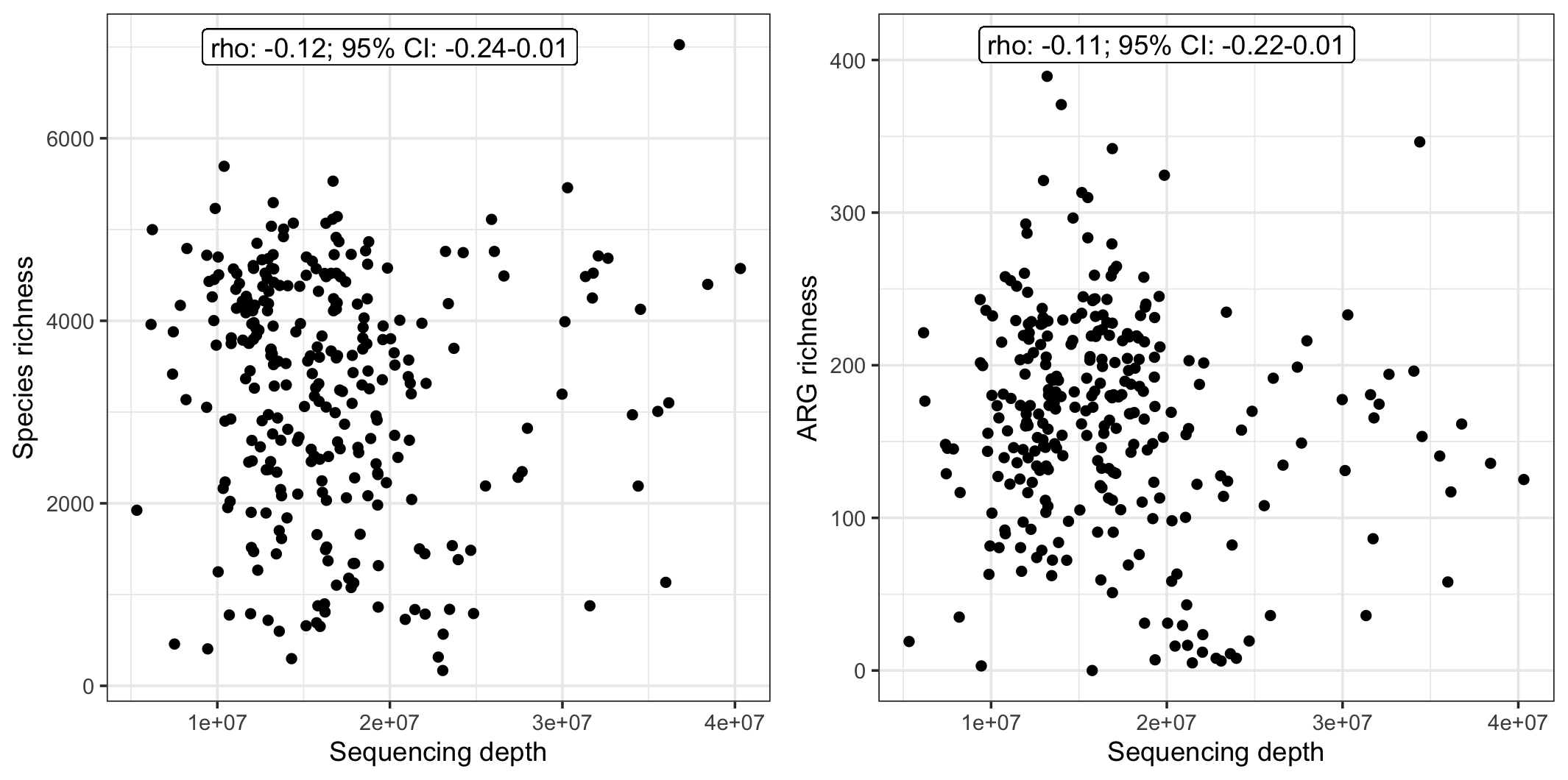

**Figure 1. Sink-trap sample species richness (left-panel) and antimicrobial resistance gene (ARG) richness (right panel) versus sequencing depth.** Panels reflect counts of unique bacterial species (left-panel) and unique ARGs identified plotted against sequencing depth. Each point represents a SinkBug sink-trap metagenome; Spearman’s rho and p-value labelled per facet.

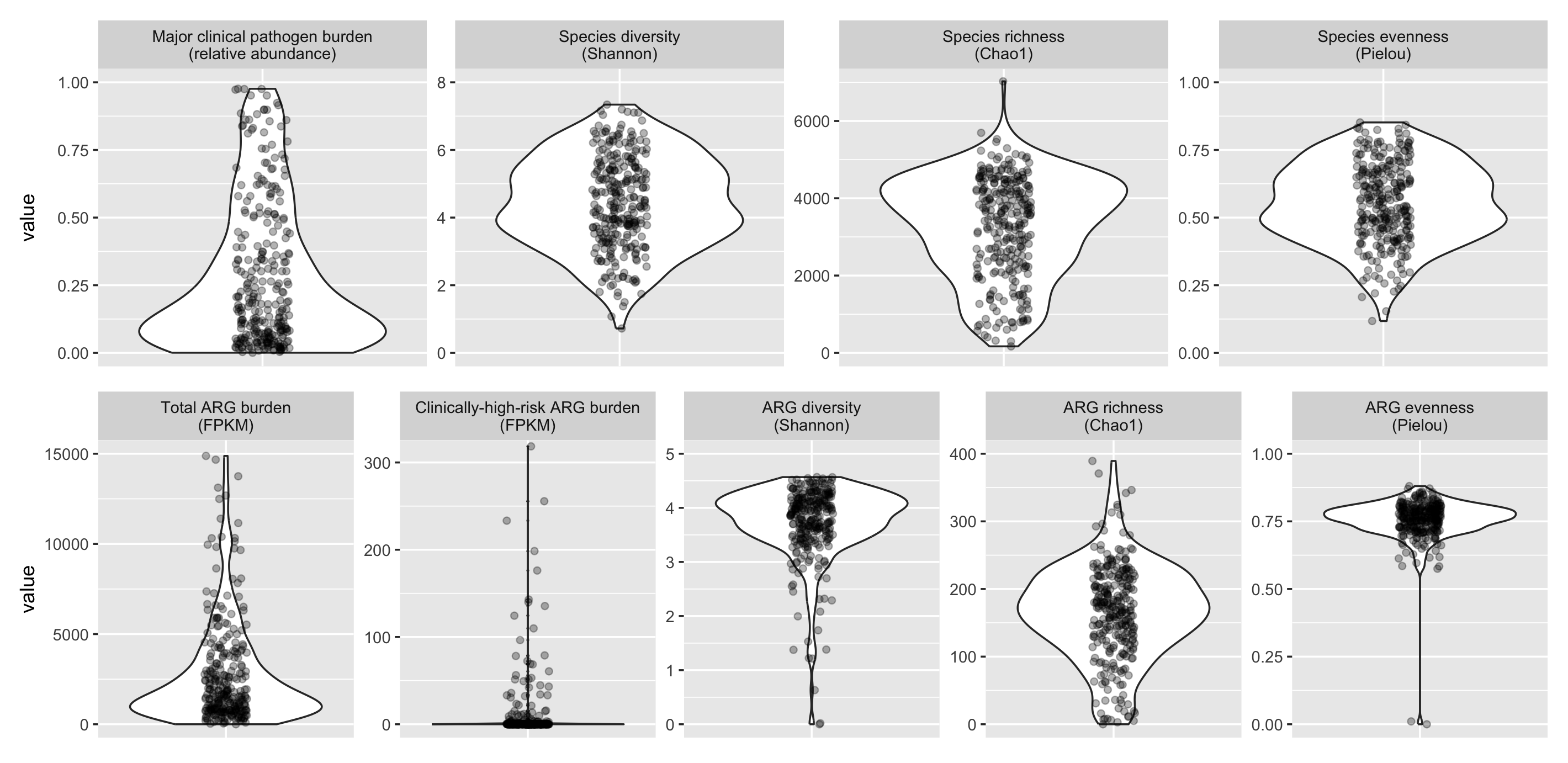

**Figure 2. Sink-trap sample composition across sink samples.** Panels reflect the distribution of nine different taxonomic and ARG composition metrics across all sink-trap samples, with each point representing a sink-trap metagenome. FPKM=Fragments per kilobase million.

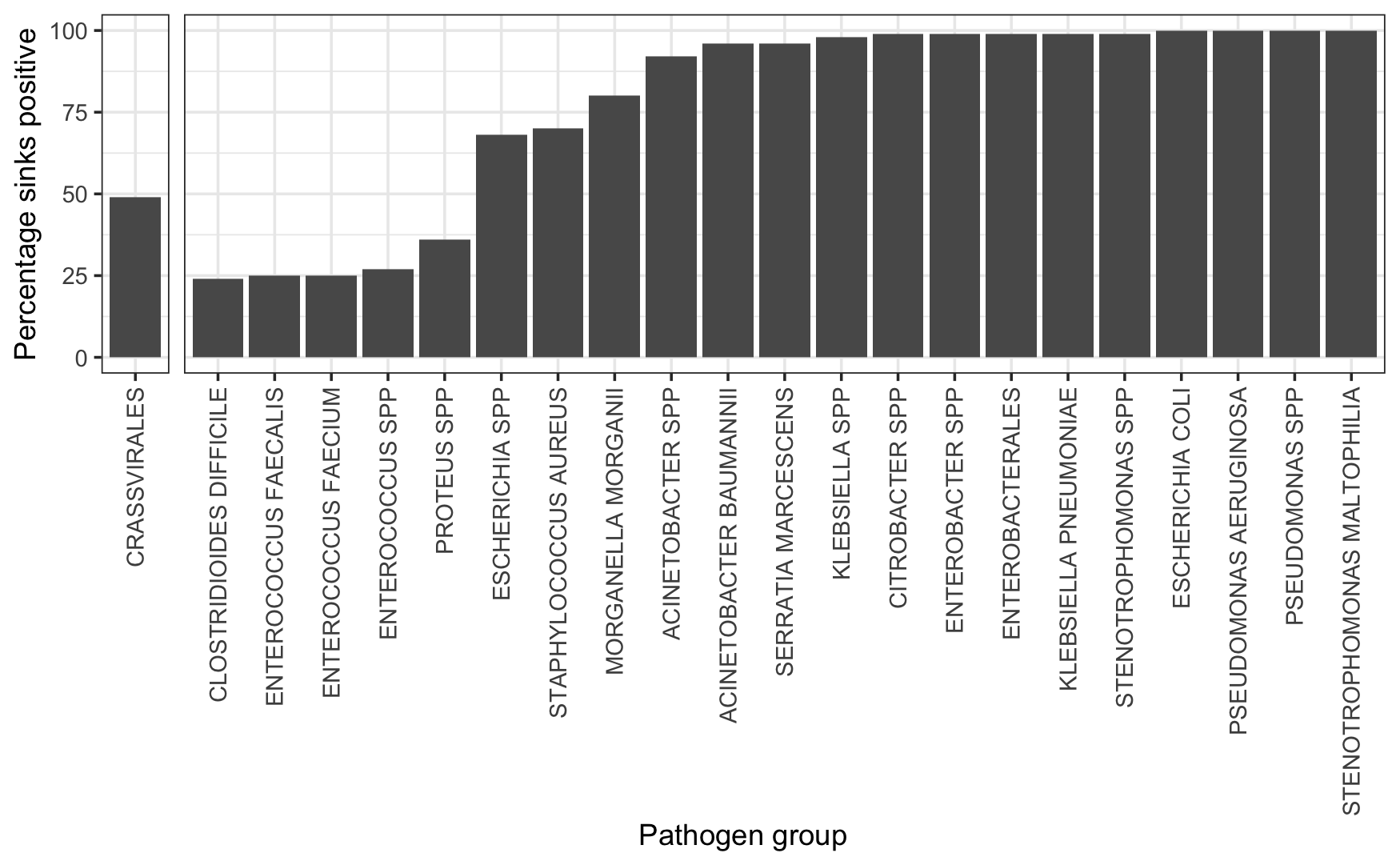

**Figure 3. Sink positivity for major clinical pathogen groups and Crassvirales.** Percentage of sink-traps (y) where any reads were identified as belonging to a major clinical pathogen group (x) or Crassvirales (consistent with faecal contamination).

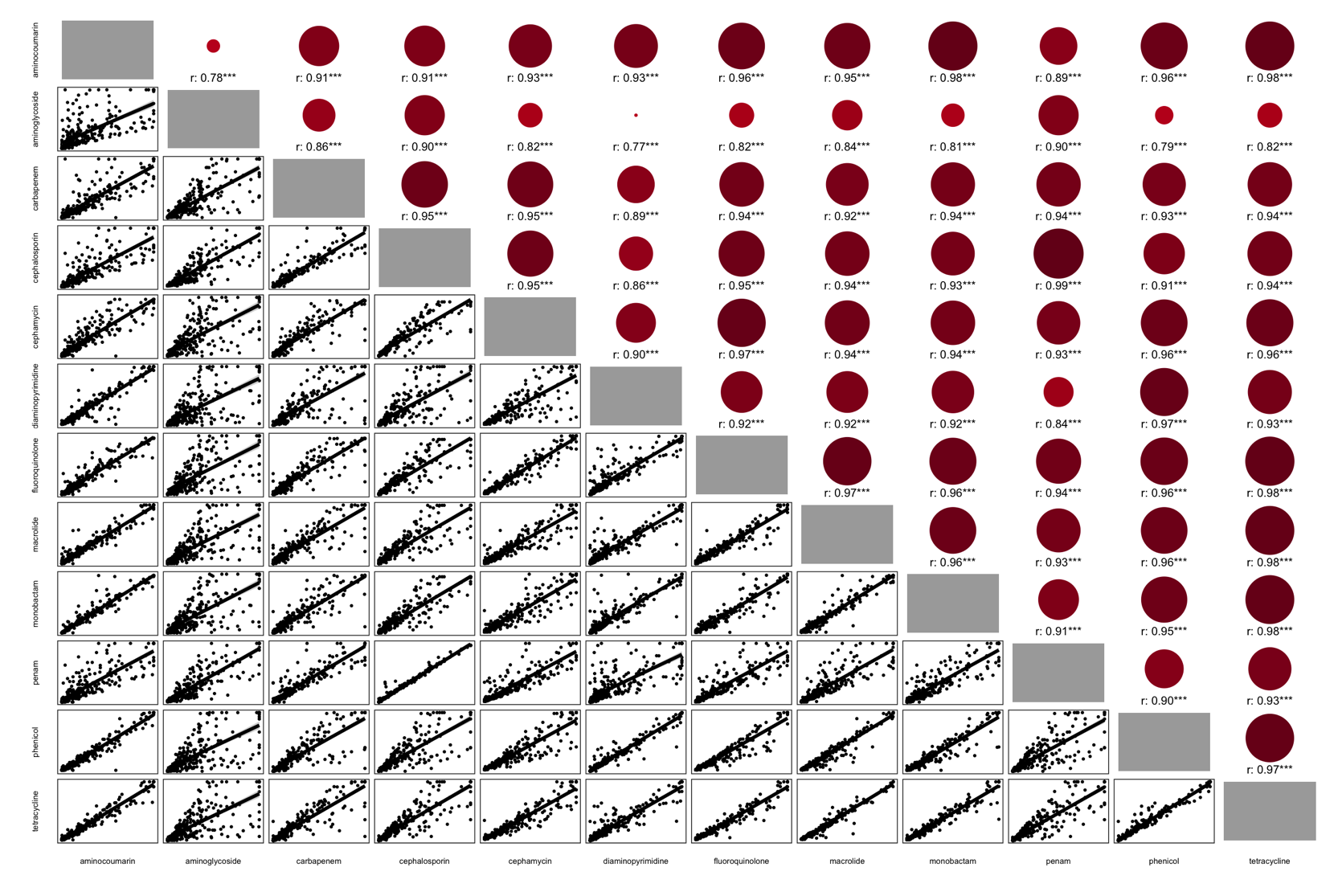

**Figure 4. Pairwise Spearman correlation matrix of antibiotic-class-level FPKM.** Correlation matrix of antibiotic-class resistance scores labelled on x and y axes per sink-trap. From bottom left: each point represents the respective antibiotic-class resistance score per sink-trap against a separate antibiotic-class resistance score for the same sink-trap; the line is a fitted “gam” smooth trend. From top right: Spearman correlations for mirrored pairwise comparisons where dot size reflects relative rho magnitude and colour reflects rho direction (red=positive, blue=negative). Spearman’s rho labelled per comparison with significance level reflected: ***<0.001; **<0.01; *<0.05; 0.1. FPKM=Fragments per kilobase million

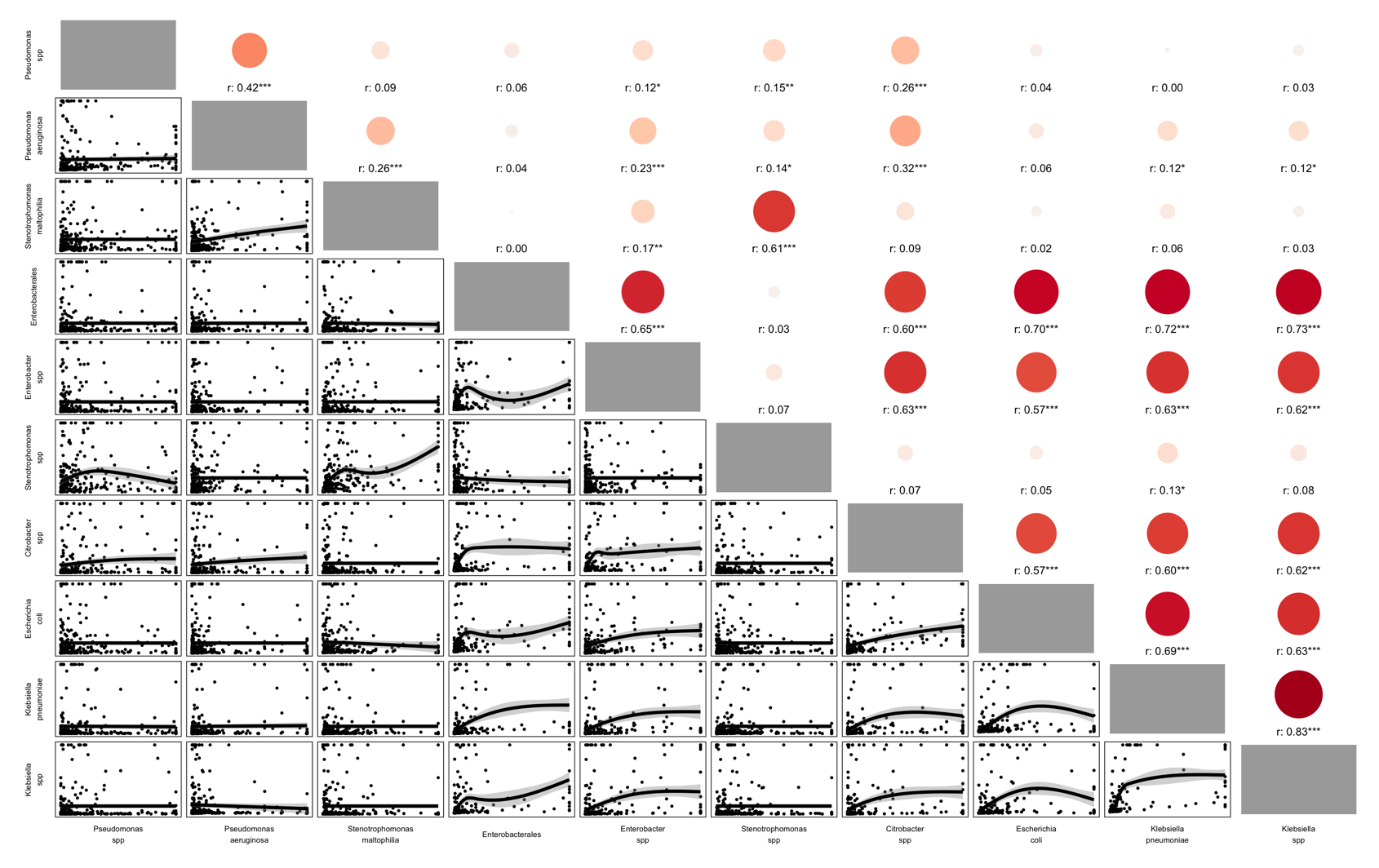

**Figure 5. Pairwise Spearman correlation matrix of major clinical pathogen abundance.** Correlation matrix of specific major clinical pathogen relative abundance labelled on x and y axes per sink-trap. Data was truncated to 5% and 95% to reduce the influence of outliers on the smoothed trends. From bottom left: each point represents the respective major clinical pathogen relative abundance per sink-trap against a separate major clinical pathogen for the same sink-trap; the line is a fitted “gam” smooth trend. From top right: Spearman correlations for mirrored pairwise comparisons where dot size reflects relative rho magnitude and colour reflects rho direction (red=positive, blue=negative). Spearman’s rho labelled per comparison with significance level reflected: ***<0.001; **<0.01; *<0.05; 0.1. Groups are exclusive, i.e. Enterobacterales is other Enterobacterales than those shown.

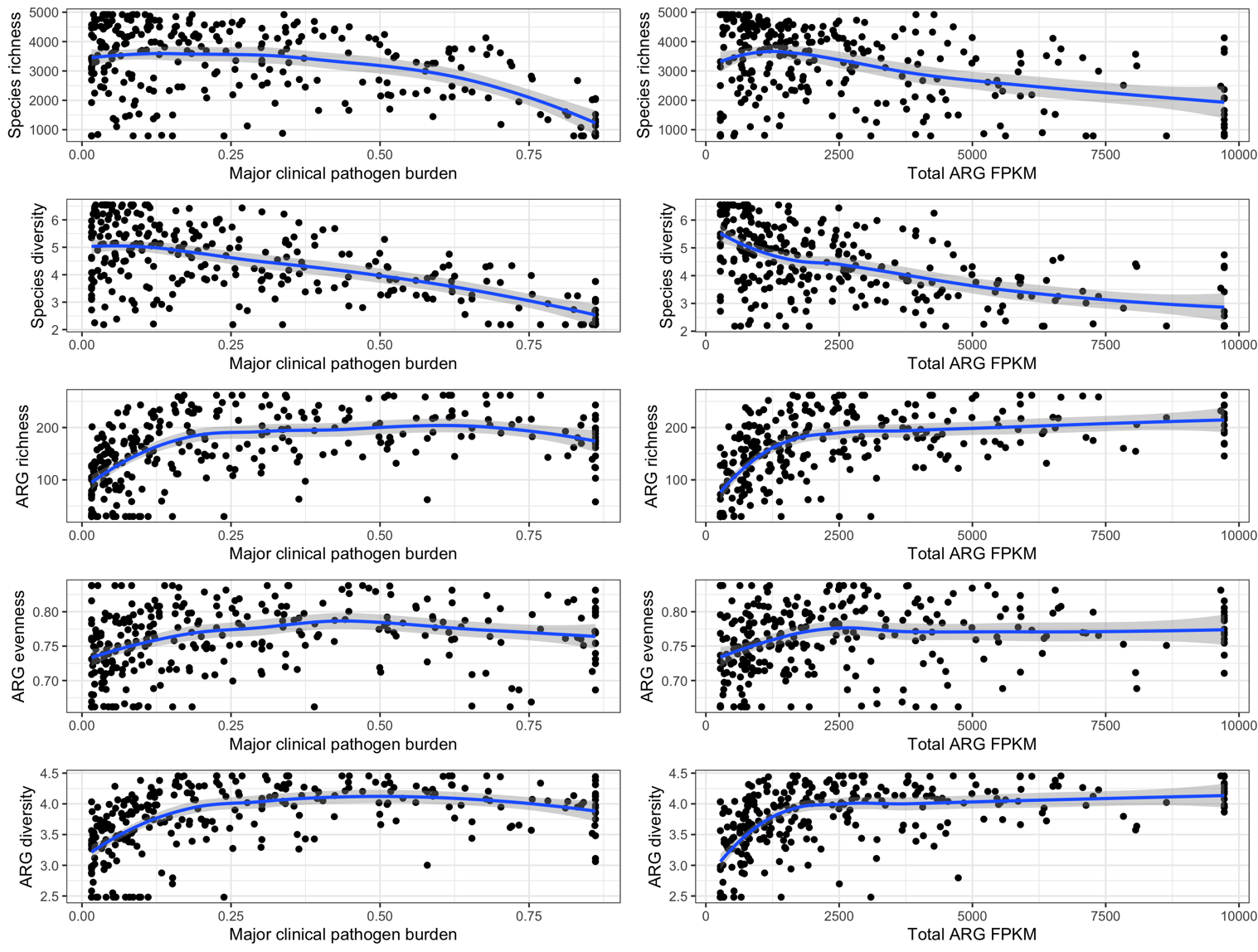

**Figure 6. Association between major clinical pathogen, total ARG burden and other diversity metrics.** Individual facets comparing pathogen/ARG burden of sink-traps against diversity metrics. Each point reflects a sink-trap and its respective burden (x) or diversity metric (y), with gam smooth trend lines and 95% confidence intervals (shaded).

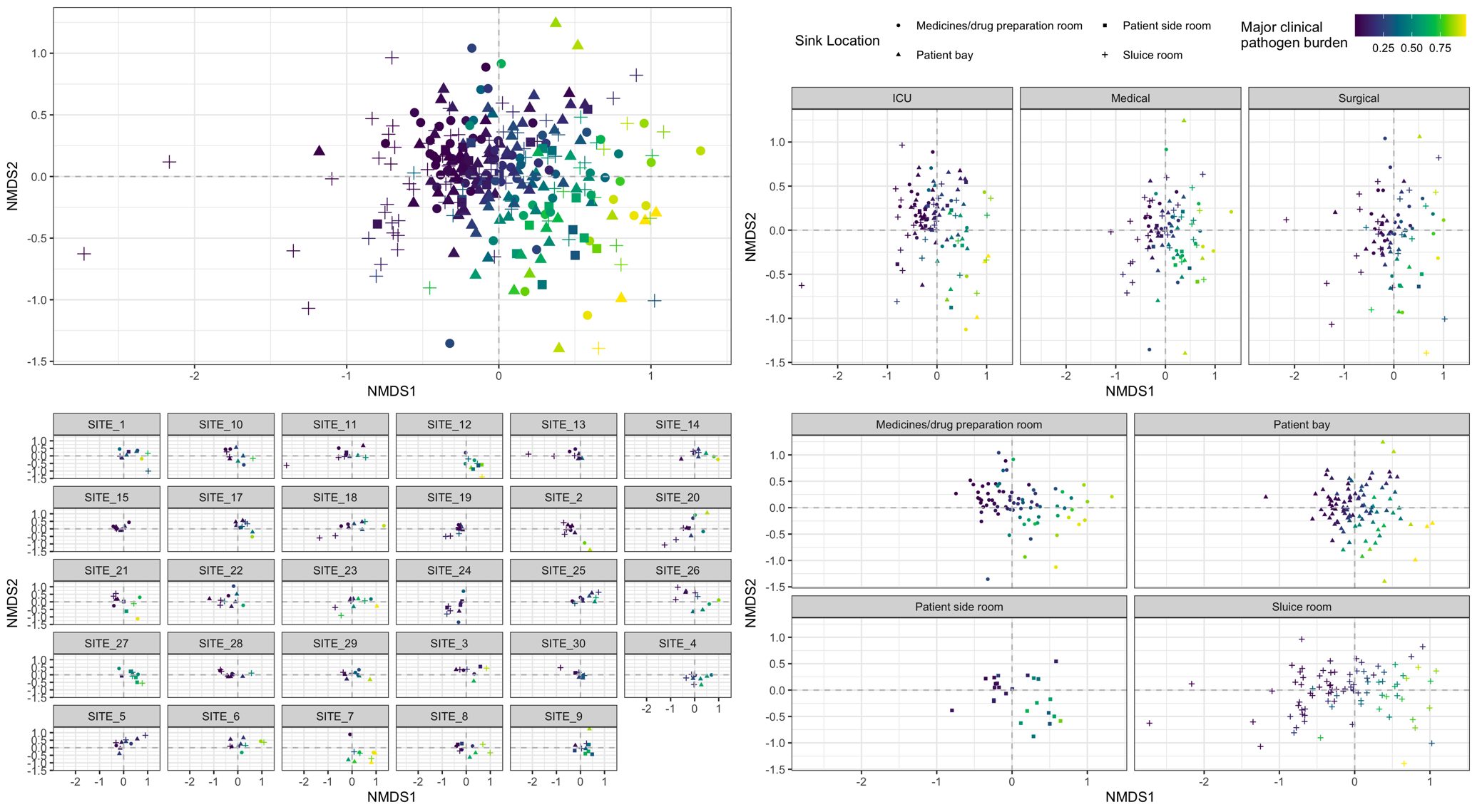

**Figure 7. NMDS ordination of sink-trap taxonomic composition.** NMDS ordination plots unfaceted (A) and faceted by ward location (B), hospital site (C) and sink location (D). Sink-traps are reflected by points where shape indicates sink location and colour indicates the total relative abundance of major clinical pathogens. Origin (0,0) is indicated by grey dashed lines.

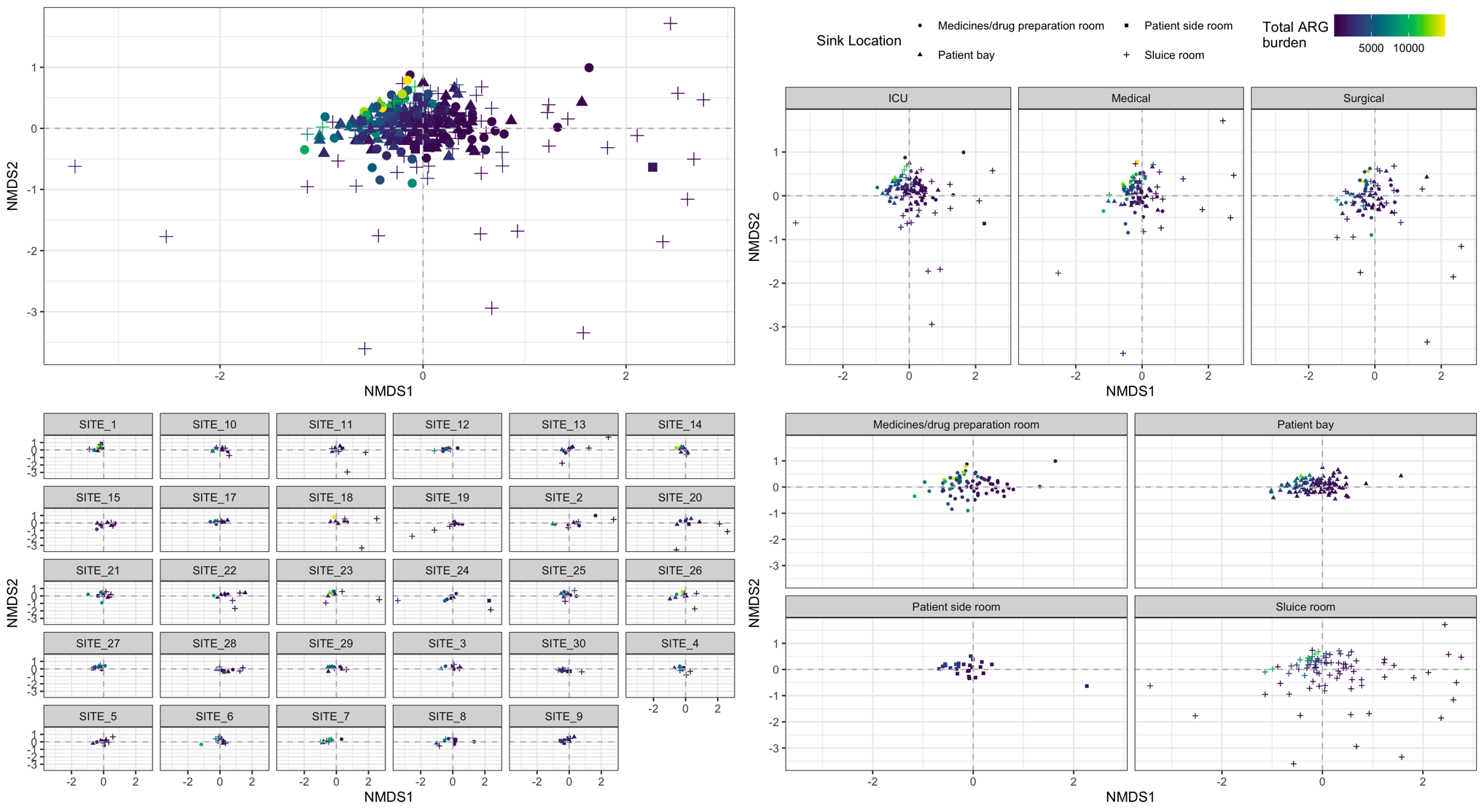

**Figure 8. NMDS ordination of sink-trap resistome composition.** NMDS ordination plots unfaceted (A) and faceted by ward location (B), hospital site (C) and sink location (D). Sink-traps are reflected by points where shape indicates sink location and colour indicates the total ARG burden (FPKM). Origin (0,0) is indicated by grey dashed lines.

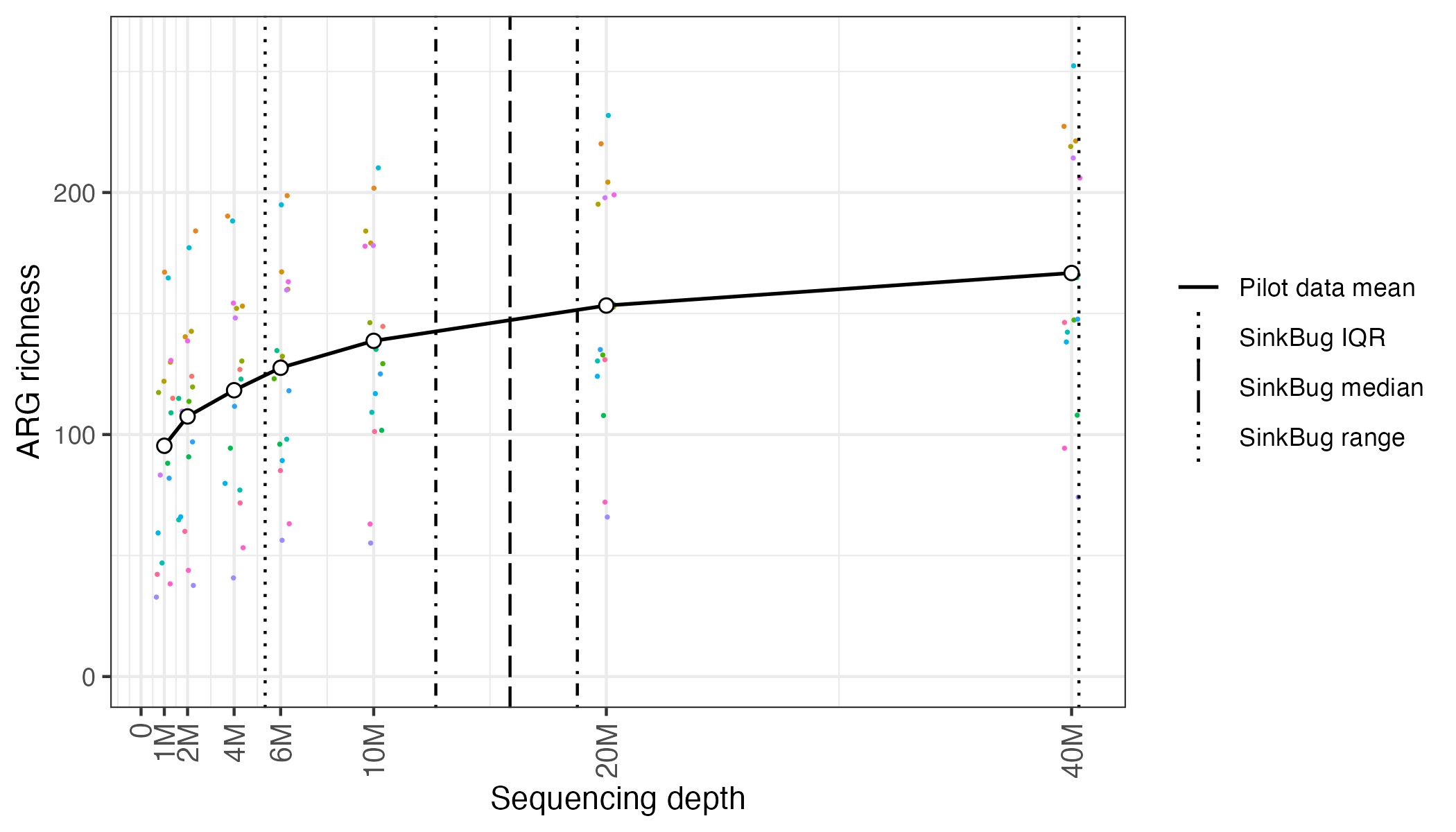

**Figure 9. Antimicrobial resistance gene (ARG) recovery in 17 previously sampled sink metagenomes subsampled to different sequencing depth.** Number of unique AMR genes (y-axis) identified in 17 previously sampled UK sink-trap metagenomes subsampled to specific sequencing depths (x-axis, number of paired-end reads) (colour reflects the same sample). Dashed line reflects median sequencing depth achieved for SinkBug (i.e. this study) metagenomes, with dot-dash and dotted lines representing interquartile range and absolute range respectively.

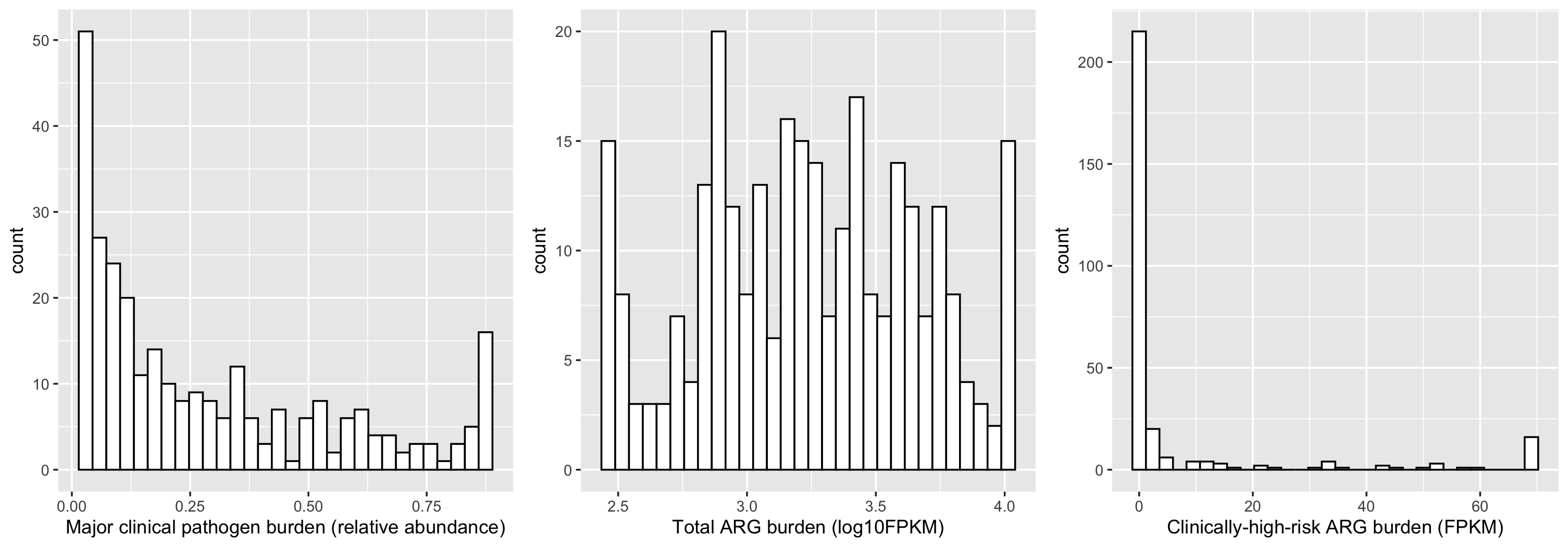

**Figure 10. Distribution of outcomes considered in the Generalised Additive Models.** Panels reflect distribution of major clinical pathogen burden, total ARG burden and clinically-high-risk ARG burden in sink-trap metagenomes, truncated to 5% and 95% percent.

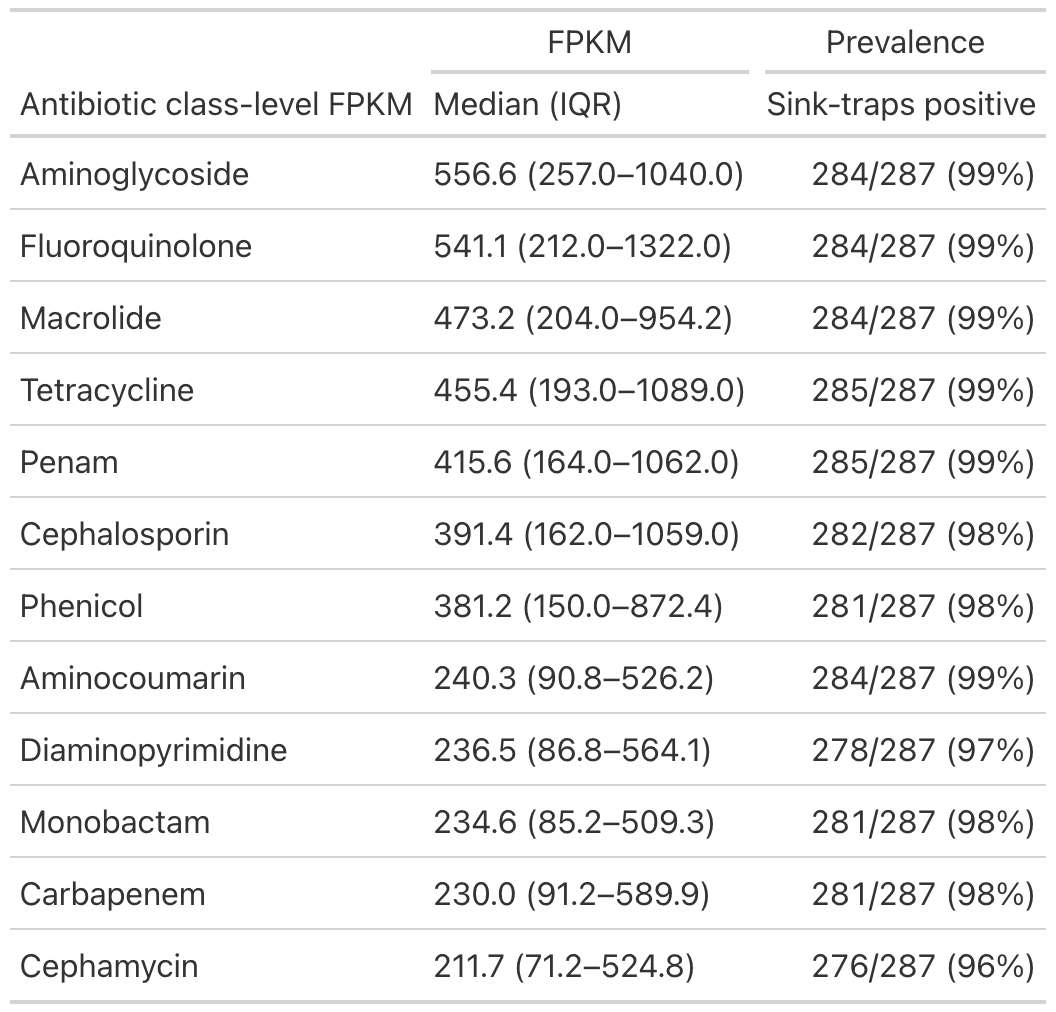

**Table S1. Antibiotic class-level resistance FPKM and sink positivity across sink-traps.**
